# GENOMIC INVESTIGATIONS OF ACUTE HEPATITIS OF UNKNOWN AETIOLOGY IN CHILDREN

**DOI:** 10.1101/2022.07.28.22277963

**Authors:** Sofia Morfopoulou, Sarah Buddle, Oscar Enrique Torres Montaguth, Laura Atkinson, José Afonso Guerra-Assunção, Nathaniel Storey, Sunando Roy, Alexander Lennon, Jack C D Lee, Rachel Williams, Charlotte A Williams, Helena Tutill, Nadua Bayzid, Luz Marina Martin Bernal, Catherine Moore, Kate Templeton, Claire Neill, Matt Holden, Rory Gunson, Samantha J Shepherd, Priyen Shah, Samantha Cooray, Marie Voice, Michael Steele, Colin Fink, Thomas E Whittaker, Giorgia Santilli, Paul Gissen, Rachel Brown, Benedikt B Kaufer, Jana Reich, Julien Andreani, Peter Simmonds, Dimah K. Alrabiah, Sergi Castellano Hereza, DIAMONDS and PERFORM consortia, Catarina Andrade, Glenn Anderson, Chayarani Kelgeri, Simon N Waddington, Juan F Antinao Diaz, James Hatcher, Surjo De, Riccardo Zenezini Chiozzi, Konstantinos Thalassinos, Thomas S Jacques, Katja Hoschler, Tiina Talts, Cristina Celma, Suam Gonzalez, Eileen Gallagher, Ruth Simmons, Conall Watson, Sema Mandal, Maria Zambon, Meera Chand, Luis Campos, Joanne Martin, Emma Thomson, Ines Ushiro-Lumb, Michael Levin, Julianne R Brown, Judith Breuer

**Affiliations:** Infection, Immunity and Inflammation Department, GOS Institute of Child Health, University College London, London, UK; Great Ormond Street Hospital for Children NHS Foundation Trust, GOSH, Department of Microbiology, Virology and Infection Control, London, UK; Genetics and Genomic Medicine Department, GOS Institute of Child Health, University College London, London, UK; Wales Specialist Virology Centre, Public Health Wales Microbiology Cardiff, University Hospital of Wales, Cardiff, UK; Department of Medical Microbiology, Royal Infirmary, Edinburgh, UK; Public Health Agency, Northern Ireland; School of Medicine, University of St Andrews, UK; West of Scotland Specialist Virology Centre; Section for Paediatrics, Department of Infectious Diseases, Faculty of Medicine, Imperial College London, London, UK; Micropathology Ltd., University of Warwick Science Park, Coventry, UK; Molecular and Cellular Immunology, GOS Institute of Child Health, University College London, London, UK; Department of Cellular Pathology - University Hospitals Birmingham NHS Foundation Trust, London, UK; Institute of Virology, Freie Universität Berlin, Berlin, Germany; Nuffield Department of Medicine, University of Oxford, Oxford, UK; Centre HospitalierUniversitaire (CHU) Grenoble – Alpes, Grenoble, 38000, France; National Center for Biotechnology, King Abdulaziz City for Science and Technology, Riyadh 11461, Saudi Arabia; UCL Genomics, University College London, London, UK; Histopathology Department, Great Ormond Street Hospital for Children NHS Foundation Trust, London, UK; Liver Unit, Birmingham Women’s and Children’s Hospital NHS Trust; Gene Transfer Technology Group, EGA-Institute for Women’s Health, University College London, London, UK; MRC Antiviral Gene Therapy Research Unit, Faculty of Health Sciences, University of the Witswatersrand, Johannesburg, South Africa; UCL Mass Spectrometry Science Technology Platform, Division of Biosciences, University College London, London, United Kingdom; Institute of Structural and Molecular Biology, Division of Biosciences, University College London, London, United Kingdom; Institute of Structural and Molecular Biology, Birkbeck College, University of London, London, United Kingdom; Developmental Biology and Cancer Department, UCL GOS Institute of Child Health and Department of Histopathology, Great Ormond Street Hospital for Children NHS Foundation Trust, London, UK; UK Health Security Agency; Centre for Genomics and Child Health, The Blizard Institute, Queen Mary University London, UK; Medical Research Council-University of Glasgow Centre for Virus Research, Glasgow,UK; NHS Blood and Transplant, UKHSA

## Abstract

Since the first reports of hepatitis of unknown aetiology occurring in UK children, over 1000 cases have been reported worldwide, including 268 cases in the UK, with the majority younger than 6 years old. Using genomic, proteomic and immunohistochemical methods, we undertook extensive investigation of 28 cases and 136 control subjects. In five cases who underwent liver transplantation, we detected high levels of adeno-associated virus 2 (AAV2) in the explanted livers. AAV2 was also detected at high levels in blood from 10/11 non-transplanted cases. Low levels of Adenovirus (HAdV) and Human Herpesvirus 6B (HHV-6B), both of which enable AAV2 lytic replication, were also found in the five explanted livers and blood from 15/17 and 6/9 respectively, of the 23 non-transplant cases tested. In contrast, AAV2 was detected at low titre in 6/100 whole bloods from child controls from cohorts with presence or absence of hepatitis and/or adenovirus infection. Our data show an association of AAV2 at high titre in blood or liver tissue, with unexplained hepatitis in children infected in the recent HAdV-F41 outbreak. We were unable to find evidence by electron microscopy, immunohistochemistry or proteomics of HAdV or AAV2 viral particles or proteins in explanted livers, suggesting that hepatic pathology is not due to direct lytic infection by either virus. The potential that AAV2, although not previously associated with disease, may, together with HAdV-F41 and/or HHV-6, be causally implicated in the outbreak of unexplained hepatitis, requires further investigation.

## Introduction

The report in March 2022 of five cases of severe hepatitis of unknown aetiology by Scottish public health authorities led to the identification by the UK Health Security Agency (UKHSA) of a total of 268 cases as of June 30^th^ 2022^1^. Cases, defined as acute non-A-E hepatitis with serum transaminases >500IU in children under ten years of age, were found to have been occurring since January 2022^2^. The incidence for acute hepatitis of unknown aetiology in children was compared to the mean number of cases over the previous five years and the 2022 incidence was at least three times higher than the five-year baseline^3^. In Europe, 473 cases have been reported as of 30th June 2022^1^, including the 268 UK cases, of which 189 required hospitalisation, including seventy-four that were admitted to the ICU and twelve which required liver transplantation^4^.

In the UK, investigations identified adenovirus as the most common pathogen with 156/241 (64.7%) testing positive in one or more samples and whole blood was found to be the most sensitive specimen for detecting the virus^5^. SARS-CoV-2 infection was detected in 17.3% of cases (34/196) although in English cases the proportion was lower (9.7%)^4^ with HHV-6 and HHV-7 detected in around 40% of the smaller number (48 and 35 respectively) of samples tested. Of the 77 cases of adenovirus (HAdV) detected in blood, 35 were found to be subtype F41 (HAdV-F41) by hexon PCR and sequencing. Seven of eight England-resident patients who required a liver transplant tested positive for adenovirus in blood, of which five were genotyped and found to be F41^2^.

Given uncertainty around the aetiology of this outbreak, and the potential that the adenovirus, if implicated (**Figure 1A**), could be a new variant or a recombinant virus, samples were sent to Great Ormond Street Hospital and University College London for genomic analyses. We undertook untargeted metagenomic and metatranscriptomic sequencing, confirmatory PCR and viral whole genome sequencing (WGS) on liver biopsies from cases requiring liver transplants (all aged ≤5 years) and on whole blood and other samples from non-transplant cases (aged <10) presenting with the unknown hepatitis (**Table 1, Figure 1B**).

**Figure 1:**
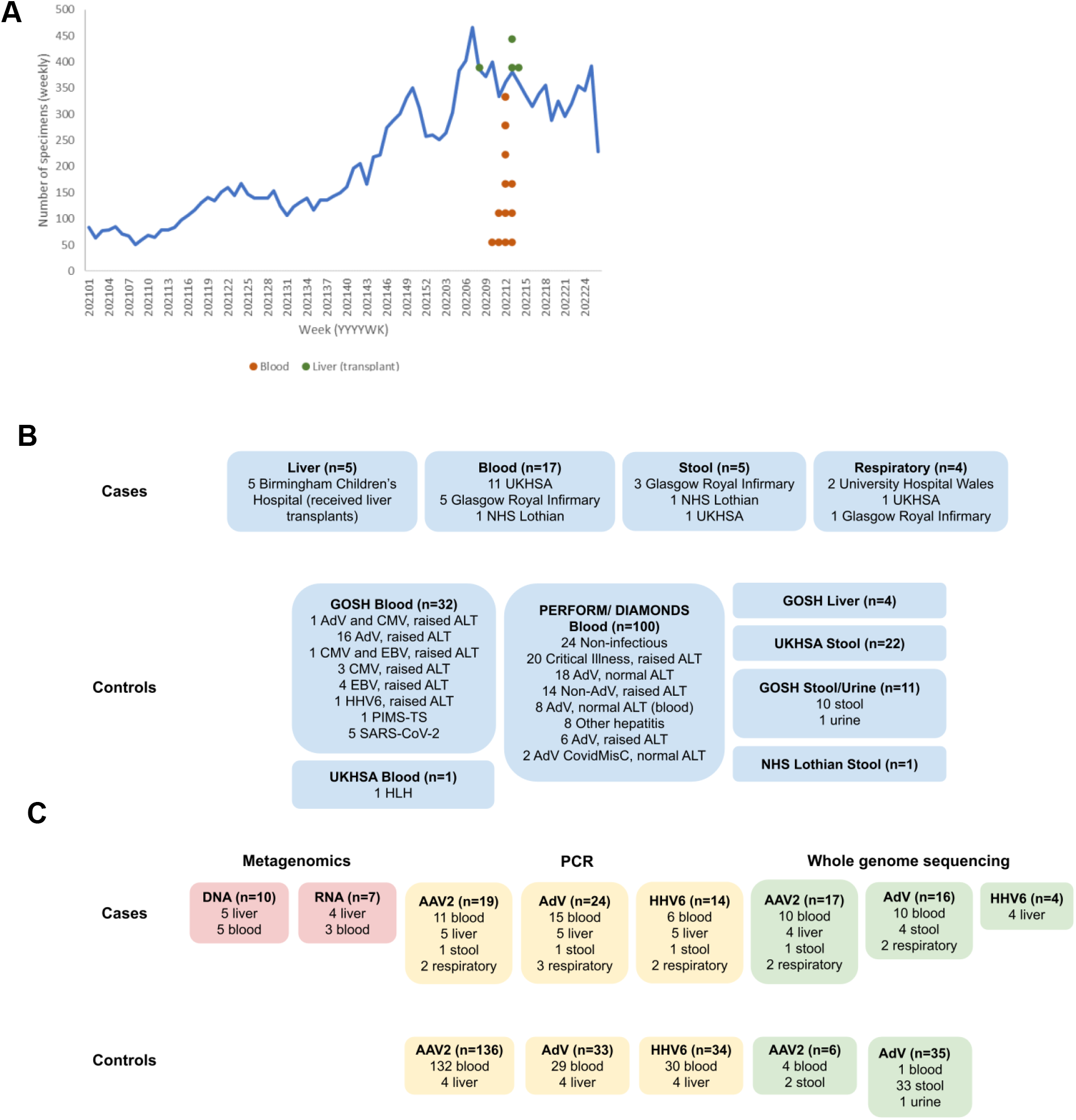
HAdV Epidemiology and experimental outline. (A) HAdV in stools; epidemiology since January 2021. Dots represent the day of presentation for the cases, in green the cases who underwent a liver transplant and in red the remaining cases, all in either late March or through April 2022. (B) Case and control specimens by source. (C) Tests carried out by specimen type. More detail on samples tested and the results can be found in Tables 1 and 2.

**Table 1:**
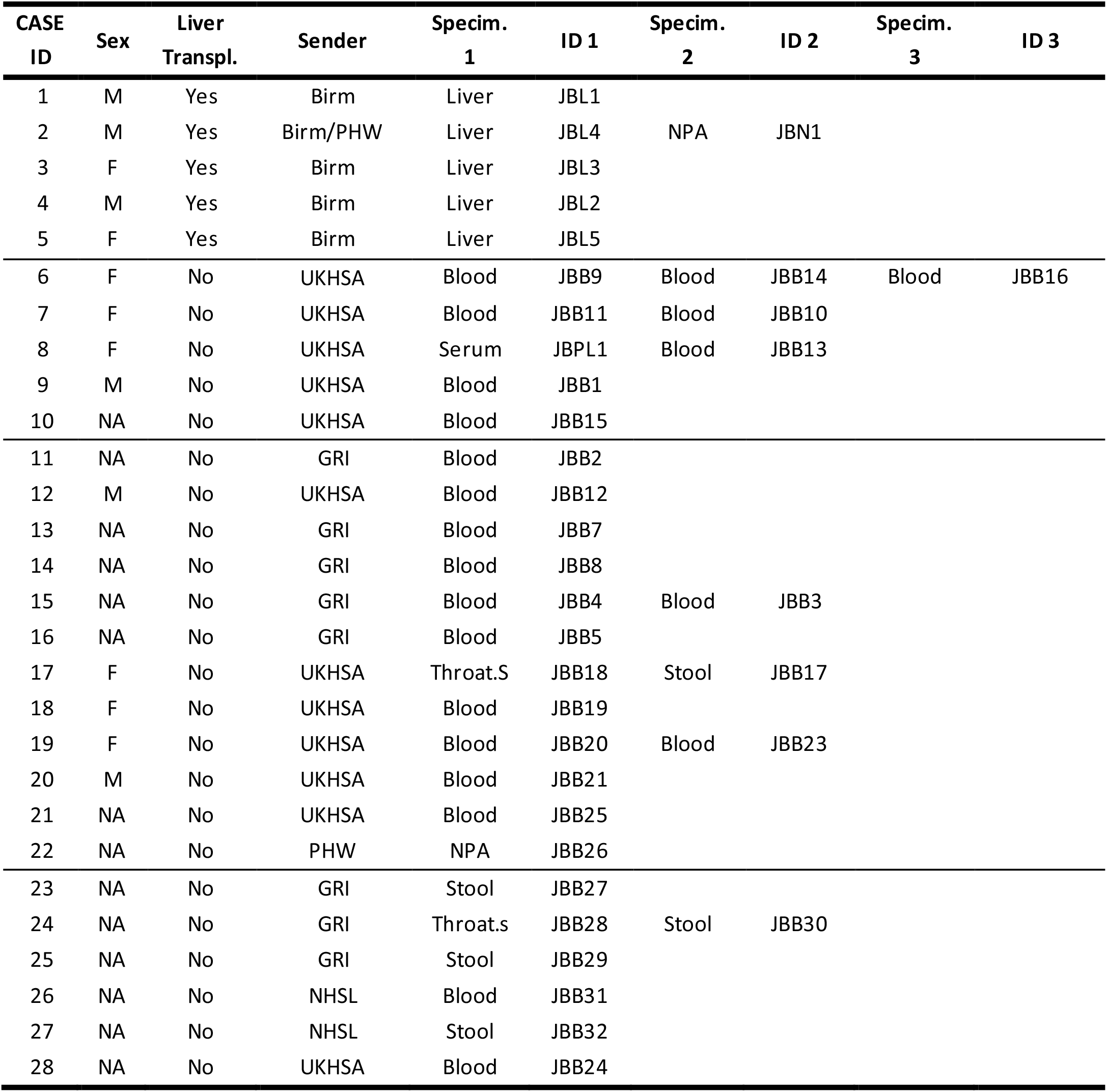
Characteristics of unexplained pediatric hepatitis cases and related specimens. The median age for the cases is 3 years old (age range: 1y-7y) **Cases 1-5** underwent liver transplant and had mNGS, PCR and WGS done on their specimens. **Cases 6-28** did not have a liver transplant. **Cases 6-10** had mNGS, PCR and WGS on their samples. **Cases 11-22** had PCR (not all viruses) and WGS. **Cases 23-28** only had HAdV WGS on their samples and there was no residual material for further testing.

To guide our understanding of the findings, we used HAdV, AAV2 and HHV-6 PCR to test residual whole blood specimens from immunocompetent and immunocompromised child controls with or without HAdV infection, including groups with raised liver transaminases (Figure 1B, Tables 3 and 4). We also analysed blood from healthy controls, patients with SARS-CoV2 and liver biopsies from immunocompromised children with non HAdV hepatitis. Immunocompetent controls (n=100) were hospitalized children, median age 5.6 years, recruited before (March 2017 - December 2019) and during the Covid-19 pandemic (June 2020-October 2021) from the PERFORM (Personalised Risk assessment in febrile illness to optimise Real-life Management,www.perform2020.org) and DIAMONDS (Diagnosis and Management of Febrile Illness using RNA Personalised Molecular Signature Diagnosis study, www.diamonds2020.eu) respectively. A second control cohort comprised 32 children, median age 2 years, with compromised immunity, predominantly following solid or bone marrow transplant and raised liver transaminases unrelated to the outbreak, who were inpatients at Great Ormond Street Hospital in 2021 and 2022. GOSH controls were viraemic, including 17 with adenovirus and raised liver transaminases. Details of the children in each of the control cohorts are given in Table 3 and Table 4.

**Table 2:** PCR, metagenomics and WGS results from cases. Repeat samples from the same case had titres within 2 CTs.

**A: cases where metagenomic sequencing was performed**
| Sample | Case | PCR CT values |  |  | Metagenomics reads |  |  |  |  |  | WGS Coverage (10X) |  |  |
| --- | --- | --- | --- | --- | --- | --- | --- | --- | --- | --- | --- | --- | --- |
|  |  | AAV2 | HAd<br>V | HHV-6 | AAV2 | DNA<br>HAd<br>V | HHV-6 | AAV2 | RNA<br>HAd<br>V | HHV-6 | AAV2 | HAd<br>V | HHV-6 |
| Liver |  |  |  |  |  |  |  |  |  |  |  |  |  |
| JBL1 | 1 | 17 | 37 | 29 | 1343 | 0 | 8 | 574 | 0 | 0 | 97 | - | 3 |
| JBL4 | 2 | 21 | 42 | 32 | 360 | 0 | 8 | 49 | 0 | 0 | 93 | - | 2 |
| JBL3 | 3 | 20 | 37 | 30 | 1189 | 0 | 4 | 95 | 0 | 0 | 98 | - | 2 |
| JBL2 | 4 | 20 | 37 | 27 | 1564 | 0 | 203 | 42 | 0 | 0 | 98 | - | 94 |
| JBL5 | 5 | 21 | 37 | 28 | 266 | 0 | 12 | F | F | F | - | - | - |
| Blood |  |  |  |  |  |  |  |  |  |  |  |  |  |
| JBB14 | 6 | 25 | P | - | 83 | 0 | 0 | 27 | 0 | 0 | 94 | F | - |
| JBB16 | 6 | 26 | 36 | 37 | 68 | 0 | 0 | 56 | 0 | 0 | 97 | F | - |
| JBB10 | 7 | 21 | 36 | 37 | 103 | 0 | 0 | F | F | F | 49 | F | - |
| JBPL1 | 8 | 25 | N | N | 277 | 0 | 0 | 165 | 0 | 0 | 94 | F | - |
| JBB1 | 9 | 19 | P | P | 1936 | 5 | 0 | 0 | 0 | 0 | 94 | F | - |
| JBB15 | 10 | N | N | 37 | 0 | 0 | 0 | F | F | F | - | F | - |

| Sample | Case | PCR CT values |  |  | WGS Coverage |  |  |
| --- | --- | --- | --- | --- | --- | --- | --- |
|  |  | AAV2 | HAdV | HHV-6 | AAV2 (10X) | HAdV (1X) | HAdV (30X) |
| Blood |  |  |  |  |  |  |  |
| JBB9 | 6 | 20.07 | 36 | 37.29 | 94 | 35.52 | - |
| JBB11 | 7 | 21.05 | 36 | 37.35 | 94 | 29.35 | - |
| JBB13 | 8 | 22.43 | N | N | 94 | - | - |
| JBB2 | 11 | 19.69 | 31 | 37.43 | 94 | 7.25 | 0.22 |
| JBB12 | 12 | 21.46 | 37 | N | 94 | - | - |
| JBB7 | 13 | 20.95 | 31 | - | 95 | - | - |
| JBB8 | 14 | 20.28 | 30 | - | 95 | - | - |
| JBB3 | 15 | 21.27 | 29 | - | 94 | 68.47 | 0.32 |
| JBB4 | 15 | 22 | 30 | - | 94 | 76.42 | 0.39 |
| JBB5 | 16 | 22.63 | 33 | - | 94 | 17.51 | 0.31 |
| JBB19 | 18 | - | 34.05 | N | - | 15.7 | - |
| JBB20 | 19 | - | 36.1 | - | - | 4.1 | - |
| JBB23 | 19 | - | 37.02 | P | - | 1.8 | - |
| JBB21 | 20 | - | 35.9 | - | - | 13.2 | - |
| JBB25 | 21 | - | 34.23 | - | - | 16.1 | - |
| JBB31 | 26 | - | P | - | - | 96.09 | 0.28 |
| JBB24 | 28 | - | 33.89 | - | - | 20.8 | - |
| Respiratory |  |  |  |  |  |  |  |
| JBN1 | 2 | 24.83 | N | N | 88 | - | - |
| JBB18 | 17 | 30.47 | 39 | 44.51 | 85 | - | - |
| JBB26 | 22 | - | 35.73 | - | - | 21.6 | - |
| JBB28 | 24 | - | P | - | - | 100 | 99.88 |
| Stool |  |  |  |  |  |  |  |
| JBB17 | 17 | 30.3 | N | N | 79 | - | - |
| JBB27 | 23 | - | P | - | - | 99.99 | 99.13 |
| JBB30 | 24 | - | P | - | - | 100 | 99.51 |
| JBB29 | 25 | - | P | - | - | 33.54 | 0.12 |
| JBB32 | 27 | - | P | - | - | 99.05/91.29 | 0.5/0.79 |

- : Not tested due to insufficient residual material,
P: Positive in referring lab
N: Negative PCR result at GOSH and/or referring laboratory

**Table 3:** GOSH controls.

| Control group | ALT (U/L) | Sample type | Number of controls |
| --- | --- | --- | --- |
| Adenovirus, raised ALT* | >500 | Blood | 16 |
| CMV, raised ALT * | >500 | Blood | 3 |
| HHV-6, raised ALT* | >500 | Blood | 1 |
| EBV, raised ALT* | >500 | Blood | 4 |
| HAdV and CMV, raised ALT* | >500 | Blood | 1 |
| CMV and EBV, raised ALT* | >500 | Blood | 1 |
| MIS-C syndrome | 103 | Blood | 1 |
| SARS-CoV2 positive* | 15–101 | Blood | 5 |
| Liver biopsy* | 198–3528 | Tissue | 4 |

\* Immunocompromised

**Table 4:** DIAMONDS and PERFORM controls (all immunocompetent)

| Control Group | PERFORM | DIAMONDS | Total |
| --- | --- | --- | --- |
| Healthy control | 24 | 0 | 24 |
| Adenovirus, normal ALT | 11 | 7 | 18 |
| Adenovirus, normal ALT (blood) | 8 | 0 | 8 |
| Adenovirus, raised ALT | 5 | 1 | 6 |
| Critical Illness, raised ALT | 9 | 11 | 20 |
| Non-adenovirus, raised ALT | 8 | 6 | 14 |
| Adenovirus CovidMisC, normal ALT | 0 | 2 | 2 |
| Other hepatitis | 7 | 1 | 8 |
| **Total** | **72** | **28** | **100** |

**Table 5:** Age distribution of control patients from GOSH, DIAMONDS and PERFORM.

| Age (years) | GOSH | DIAMONDS | PERFORM |
| --- | --- | --- | --- |
| <1 | 7 | 3 | 8 |
| 1-5 | 16 | 11 | 33 |
| 6-10 | 3 | 5 | 14 |
| 11-17 | 6 | 9 | 17 |
| **Total** | **32** | **28** | **72** |

## Results

### Metagenomic Sequencing

We performed metagenomic and metatranscriptomic sequencing on 5 liver samples from 5 transplant cases and 6 blood samples from 5 non-transplant cases (**Table 1, Figure 1B**). The 5 liver samples had uniform and consistently high sequencing depth both for DNA-seq and RNA-seq (**Supplementary Table 2**) while the 6 blood samples had more variable sequencing depth particularly for RNA-seq (**Supplementary Table 2**). Reads from adeno-associated virus 2 (AAV2), a member of the *Dependoparvovirus* genus were the most abundant in both DNA-seq (7-42 read per million) and RNA-Seq (0.7-10 reads per million) from all five explanted liver biopsy samples (**Table 2A**). Lower AAV2 read counts (1.2-42 reads per million) were detected in blood from 4/5 non-transplant cases (**Table 2A**). We detected lower levels of human herpesvirus 6B (HHV-6B) in the DNA-Seq of all five explanted liver samples (0.09-4 reads per million) but not in the RNA-seq or the 6 blood samples (**Table 2A**). HAdV was not detected in liver and was present (5 reads) in only one blood sample (**Table 2A**).

### Confirmatory real-time PCR

#### Cases

We tested a total of seventeen cases for AAV2 by PCR, including five transplant cases and twelve non-transplanted cases.

PCR for AAV2 was positive in all five explanted livers at low cycle threshold (CT) values (CT range: 17-21) indicating high viral titres (**Table 2, Figure 2A**). HHV-6 and HAdV were also detected in all the liver samples, CT ranges: 27-32 and 36-41 respectively.

**Figure 2:**
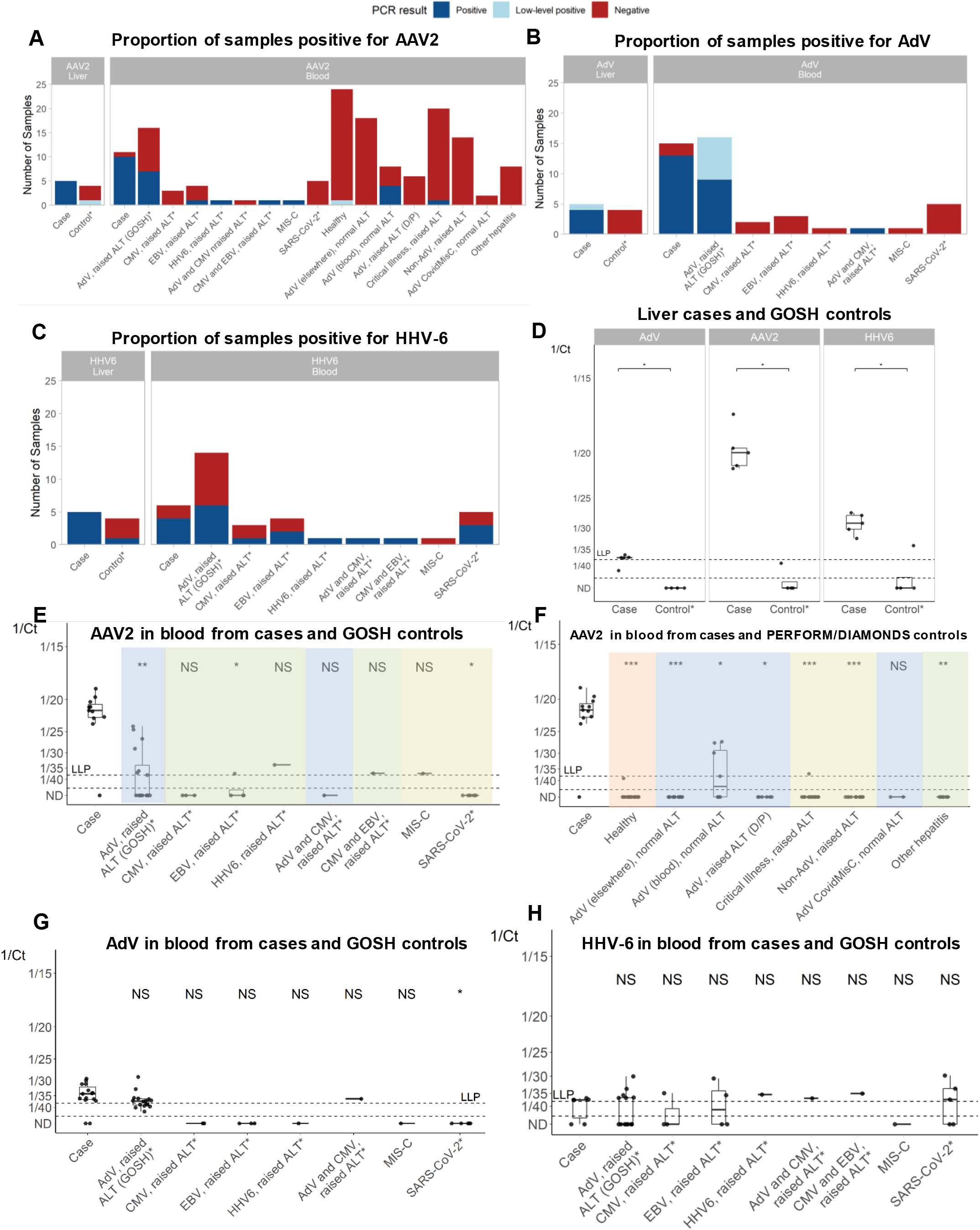
viral loads (CT values) for cases and controls. * indicates immunocompromised controls Proportion of PCR positive and negative results for A) AAV2, B) AdV and C) HHV-6. Ct values of below 38 were defined as positive. Ct values of above 38 but where the virus was detected within the maximum 45 cycles were defined as low-level positive. D) AdV, AAV2 and HHV-6 in liver cases and immunocompromised GOSH liver controls. E) AAV2 in blood from cases and GOSH controls (all immunocompromised apart from MIS-C patient). F) AAV2 in blood from cases and PERFORM/DIAMOND controls. Blue: AdV infection, green: non-AdV hepatitis, red: healthy, yellow: other controls. G) AdV levels in blood from cases and immunocompromised GOSH controls. H) HHV-6 in blood from cases and immunocompromised GOSH controls. In the box plots, the bold middle line represents the median and the upper and lower horizontal lines represent the upper and lower quartiles respectively. Each point represents one case or control. Where more than one sample for a case was tested, the midpoint of the CT values is shown (all repeats were identical ie≤2CT values -Table 2). The dotted line marked LLP indicates the low-level positive threshold (38). Points below the second dotted line represent samples below the limit of PCR detection (45). Wilcoxon non-parametric rank sum tests were conducted for (D), and a Kruskal-Wallis test followed by pairwise Wilcoxon tests for (E-H). NS: not significant, * p < 0.05, ** p < 0.01, *** p < 0.001. The single serum sample was not included in the analysis.

21/23 of non-transplant cases tested at GOSH and/or the referring laboratory, were positive by PCR for HAdV, including 15/17 in blood. The majority were genotype HAdV-F41. Material was available from twelve non-transplant cases for further testing in different assays, including five cases whose blood had already been analysed by metagenomic methods. In total, blood was tested in eleven cases by PCR for AAV2, with 10/11 cases testing positive (**Figure 1B, Table 2A, 2B**). The twelfth case was also positive for AAV2 in a throat swab and stool (**Table 2B**). Residual blood from 9/12 non-transplant cases was available for HHV-6 testing and six were positive at high CT values. Including the livers and blood, 11/14 cases were positive for HHV-6. One non-transplant case was negative by metagenomics and PCR for both HAdV and AAV2 but positive for HHV-6. While this case met the case definition ie aged <10 years with non-A-E hepatitis, there was no further information on what other investigations had been carried out to exclude other possible causes of hepatitis.

### Comparison of cases with controls

#### Liver biopsies

Samples from non-aged matched paediatric controls aged <16 years with adenovirus and/or raised liver transaminases were tested for AAV2 and HHV-6 (**Table 3**). Four residual control liver biopsies from immunocompromised children sent to the clinical microbiology laboratory at GOSH for routine investigation, were also tested (**Figure 1B, Table 3**). Three of these control patients had elevated liver enzymes with information missing for one. The median age of controls was ten years compared with three years for the cases. HAdV was not detected in any of the control liver samples, while AAV2 was positive in one with a CT value of 39. (**Figure 2D**). HHV-6B was detected in 1/4 control livers with similar CT values to the cases (**Figure 2D**).

#### Whole blood samples

Blood from 32 GOSH paediatric control patients, including 26 immunosuppressed sampled in 2021-2022, with raised liver transaminases (AST/ALT>500IU) and viraemia (adenovirus, HHV-6, CMV or EBV) were tested for AAV2 and HHV-6B (**Figure 1B, Supplementary Table 3**). The majority had received human stem cell or solid organ transplants, and none were linked to the recent hepatitis outbreak. We also tested a further five whole bloods from GOSH immunocompromised controls with SARS-CoV-2 positive respiratory specimens and one from a patient with Multisystem Inflammatory Syndrome in Children (MIS-C) syndrome (**Table 3**). One blood sample received from UKHSA from a patient with adenovirus that had raised liver enzymes, who did not meet the case definition, was also tested. In addition, 100 blood samples from immunocompetent paediatric patients from the DIAMONDS and PERFORM studies (**Table 4**), of known adenovirus infection status, with or without hepatitis, including some who were critically ill, as well as healthy controls, were also tested by PCR for AAV2 (**Figure 1B, Supplementary Table 3**).

10/11 (91%) of cases who were not transplanted were AAV2 positive in blood by PCR, compared with 6/100 (6%) and 11/32 (34%) of blood samples from DIAMONDS/PERFORM and GOSH control cohorts respectively **(Figure 2A**, p=1.617e-09 and p= 0.001533 respectively, Fisher’s exact test), with AAV2 levels being significantly higher in cases than in any of the controls (**Figure 2E and F**, p values=1.04e-15 and 4.113e-06 respectively, Mann-Whitney Test). Of the AAV2 positive GOSH controls 7/11 and 1/11 were positive for HAdV and HHV-6 respectively while 4/6 of the AAV2 positive from DIAMONDS/PERFORM were in an HAdV positive group (**Figure 2F Supplementary Table 3**). Notably, the highest AAV2 levels in controls occurred in the GOSH immunosuppressed cohort with HAdV viraemia and hepatitis (7/17 AAV2 positive, 41%) but still at statistically lower levels than in cases (p= 0.00160) (**Figure 2E**). DIAMONDS / PERFORM immunocompetent controls with HAdV infection with or without raised liver enzymes (**Figure 2F**) were infrequently AAV2 positive (4/32) and at much lower levels than the cases (p=5.7e-06). Additionally, the UKHSA whole blood sample from a patient with adenovirus with raised liver enzymes who did not meet the case definition was negative for AAV2 despite being positive for the outbreak strain of HAdV-F41. The proportion of cases positive for HHV-6 and their viral loads were similar to GOSH controls (**Figure 2C and H**). The above findings do not change when comparisons were restricted to cases and controls <10y (**Supplementary Figure 1**).

#### Evidence of AAV2 replication

The metatranscriptomic data revealed high levels of AAV2 RNA reads with no RNA detected for HHV-6 or HAdV. Mapping of the liver RNA-seq to the RefSeq AAV2 genome (NC_001401.2) identified high expression of the cap ORF, particularly at the 3’ end of the capsid, a pattern suggesting viral replication^6^ (**Supplementary Figure 3A**), while in the blood RNA reads mapped throughout the genome, albeit at much lower levels (**Supplementary Figure 3B**). Data from RT-PCR confirmed the presence of mRNA from the cap ORF in 2/4 of the liver samples (**Supplementary Table 7**).

#### Immunohistochemistry and Electron Microscopy

By immunohistochemistry, all four liver explants were negative for adenovirus with antibody clones 2/6 and 20/11, which detect all adenovirus genotypes. Staining with AAV2 mouse monoclonal antibody (clone B1) and AAV2 rabbit polyclonal antibody demonstrated some positive staining material having the appearance of non-specific ingested debris and no nuclear staining was observed (**Supplementary Figure 4**). The same pattern was also demonstrated with AAV2 antibody clones A1 and A69. Nuclear staining was observed for AAV2 infected cell lines and murine infected tissue (scattered cells only).

Liver sections were morphologically suboptimal for electron microscopy, but no viral particles were identified in hepatocytes, blood vessel endothelial cells and Kupffer cells.

#### Proteomics

We carried out proteomics analysis of livers. The results showed no evidence of AAV2, HAdV, HHV-6 viral proteins present in the explanted livers, while Class II HLA alleles were highly abundant (**Supplementary Figure 5**).

#### Whole genome sequencing

We undertook HAdV WGS for the 12 of the non-transplant cases investigated above and an additional 11 cases referred by public health agencies for HAdV WGS. There were insufficient levels of HAdV in the livers to obtain WGS. We also sequenced HAdV from controls, including the control with adenovirus and raised liver enzymes referred from UKHSA. Maximum Likelihood (ML) phylogeny of HAdV-F41 whole genome sequences (**Supplementary Table 4**) showed clustering of sequences obtained from one stool from a case with UK, with HAdV-F41 sequences collected from children at GOSH in 2020 and with the contemporary non-case with HAdV-F41 and raised liver enzymes (**Figure 3A**). There was no evidence of recombination with other adenoviruses. Whole genomes were not obtained from the blood of any case due to high CT values, however partial sequences were identified as HAdV-F41 with reads positioned across the entire viral genome, again excluding a recombinant virus. Comparison with other HAdV-F41 genomes did not identify any SNPs unique to the one HAdV-F41 genome from a case. Given reported mutation rates for HAdV-F41 and other adenoviruses^7,8^, the SNP differences between the case and other HAdV-F41 sequences are likely to have arisen before the outbreak. Importantly, compared with RefSeq sequences, no new or unique amino acid substitutions were noted in the E1a, E2a and E4 HAdV-F41 proteins, which are known to be critical for AAV2 replication^9^.

**Figure 3:**
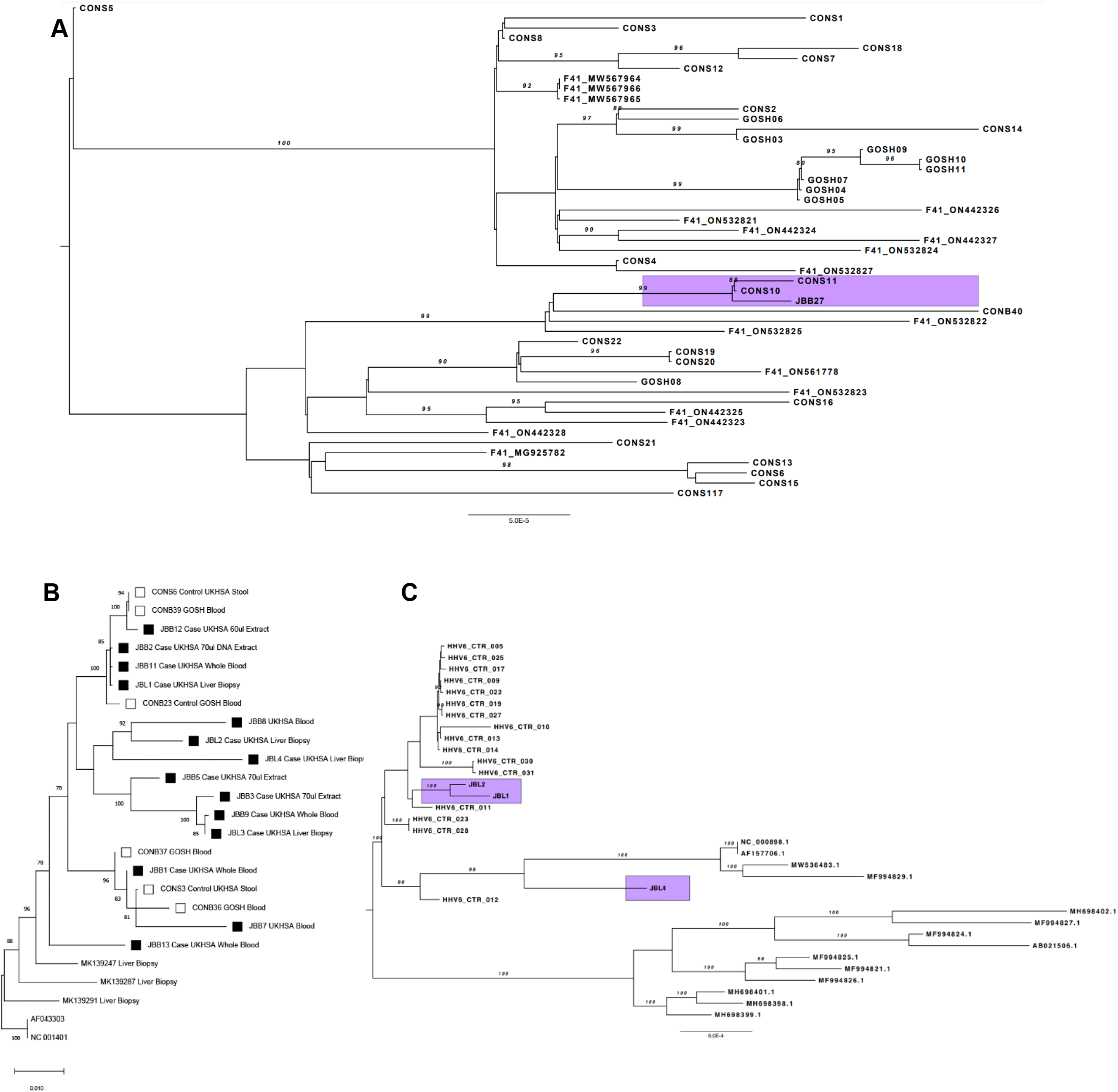
Phylogenetic trees of AdV, AAV2 and HHV-6. Maximum likelihood phylogenetic trees combining reference sequences retrieved from the RefSeq database, publicly available complete genomes from GenBank, UK non-outbreak controls as well as sequences from cases that are part of the outbreak under investigation (highlighted) for the different viruses involved. A) AdV B) AAV2 and C) HHV-6

Maximum likelihood phylogeny showed clustering of AAV2 sequence from fourteen cases, including four livers and eleven bloods from non-transplanted cases, with sequences from six control samples positive for AAV2 (**Figure 3B, Supplementary Table 5**). The degree of diversity and lack of a unique common ancestor between genomes from cases suggests that they are not specific to this outbreak, but instead reflect the current viral diversity in the general population. Some AAVs are known to be hepatotropic including AAV7 and AAV8. Comparison of the AAV2 sequences show no difference between cases and controls. However contemporary AAV2s showed changes in the capsid compared to historic AAV2. None of these changes were shared with the hepatotropic AAV (**Supplementary Figure 2**). While mean read depths for four HHV-6 genomes recovered from explanted livers were low (x5-x10) (**Supplementary Table 6**), phylogeny (**Figure 3C**) confirmed that all were different, excluding the possibility of contamination.

#### *De novo* assembly of unclassified reads

To exclude the possibility of a novel pathogen with insufficient similarity to the pathogens in the reference databases we use for taxonomic classification, we *de novo* assembled the unclassified reads from the metagenomics datasets. All of the contigs longer than 1000bp matched to human except two which mapped to Torque Teno virus (TTV).

## Discussion

Since the first reports of hepatitis of unknown aetiology occurring in UK children, over 920 cases have been reported worldwide with at least 45 (5%) requiring liver transplantation and 18 (2%) reported deaths^10^. In the UK, the majority of the 268 cases have been under the age of 6 years with at least 39% (74/189 cases) of hospitalised cases requiring admission to intensive care. Where testing is available, most cases have been positive by hexon typing methods for adenovirus (n=142/216) and specifically for HAdV-F41 (n=27/35) in blood^10^, a finding also confirmed in two cases series from the USA and UK^11^,^12^. HAdV-F41 is normally associated with benign if unpleasant gastroenteritis in children and has not been reported to cause hepatitis. HAdV-F41 viraemia, present in the majority of cases of unknown hepatitis, is recognized in immunocompromised children^13,14^ and has also been reported in healthy children with diarrhoea^15^ who were sampled opportunistically. These findings suggest that the absence of previous reports of HAdV-F41 in blood may represent under sampling. We found no evidence for a change in circulating HAdV-F41 genomes to explain the cases of hepatitis with no viral recombination detected and close clustering of the single genome from a case with non-outbreak HAdV-F41 genomes and one control HAdV-F41 genome sampled during the outbreak (**Figure 3A**). Metagenomic DNA sequencing of samples from cases detected HAdV-F41 in only one of five blood samples and none of five liver samples. However, the methodology used by us is known to be up to 2 logs less sensitive than PCR for detection of pathogen DNA in tissue and indeed PCR detected HAdV-F41, albeit at very low levels in all livers (**Table 2A**). The viral CT values in blood from cases were also low (**Table 2, Figure 2E**).

In contrast AAV2, a member of the *Parvoviridae*family was highly abundant both metagenomically and by PCR both in the five explanted livers and in blood samples from 11/12 non-transplanted cases, but detected infrequently in GOSH, DIAMONDS and PERFORM controls (**Figure 2B**). Replication of AAV2 requires coinfection with a helper virus, such as HAdV, herpesviruses, or papillomavirus^16^ and almost all the low level AAV2 detected in controls was in association with HAdV or HHV-6 infection, albeit at lower levels than in the cases (**Figure 2F,2G**). Despite AAV2 establishing latency in liver^17^, hepatitis per se, particularly in the immunocompetent DIAMONDS and PERFORM controls, was not necessarily associated either with detection or high levels of AAV2 (Figure 2G), refuting the likelihood that the AAV2 levels seen in cases resulted from reactivation as a result of liver damage. Neither have we identified AAV2 as an incidental finding in independent metagenomic studies of blood (n=350) from febrile children and SARS CoV-2 positive adult patients or livers (n=3) from patients sampled prior to the COVID-19 pandemic (Morfopoulou et al., Breuer et al unpublished data). Intriguingly, the highest AAV2 levels in control samples occurred in immunocompromised (GOSH) controls with HAdV viraemia and hepatitis (**Figure 2F**), while levels in the DIAMONDS and PERFORM controls with HAdV infection were lower (**Figure 2G)**. The higher levels of AAV2 in the GOSH controls with adenoviraemia and hepatitis as compared with other HAdV infected controls could provide further support for a specific role for AAV2 in HAdV hepatitis. However, as we do not know whether hepatitis in these controls was caused by adenovirus and other explanations are also possible, this needs further investigation. HHV-6, although present in all liver explants, was not detected more commonly or at higher titre in the blood of cases compared to controls.

RNA transcriptomic and rt-PCR evidence of replication in explanted livers point to active AAV2 infection. However, we did not detect any AAV2, HAdV, or HHV-6 proteins by IHC (**Supplementary Figure 4**) or proteomics (**Supplementary Figure 5**). Nor were viral particles visible on electron microscopy. It is unlikely that HAdV-F41 titres had declined in the interval between the onset of viral insult and clinical presentation, as the five transplanted children for whom we had data, had a median of only 12 days (**Supplementary Table 1**) from first symptom onset to transplant. The AAV2 REP78 protein is known to bind to and downregulate adenovirus DNA replication^16^, which may explain the low HAdVtitres in affected cases, although not the absence of AAV2 proteins. Downregulation of HAdV helper proteins has been shown to promote the establishment of AAV2 latency in multiple cells types^18,19^, with wild-type AAV2 largely persisting episomally^18^. More rarely, AAV2 is capable of chromosomal integration^20^, either into a specific site in chromosome 19^21^ but also throughout the genome at lower frequency^22^. Recombinant AAV gene therapy vectors which contain only the inverted terminal repeats associated with genomic integration, but no other viral sequences, have not generally been associated with hepatitis. However, liver dysfunction following gene therapy with an AAV8 vector was reported in two boys with X-Linked Myotubular Myopathy^23^ and attributed to the high doses of the vector administered and possibly to pre-existing liver pathology. AAV8 is known to be more hepatotropic than AAV2, and this property has been associated with particular capsid residues^24^,^25^. While we did not find in silico evidence that changes in recently circulating AAV2s are likely to be hepatotropic, this needs further investigation (**Supplementary Figure 2B**).

Increased circulation of the SARS-CoV-2 Omicron variant preceded the outbreak of unknown hepatitis. Three of the five transplanted cases were positive at presentation for SARS-CoV-2 by PCR and one of five had positive serology (**Supplementary Table 1**), Seven of eight non-transplanted cases tested serologically for SARS-CoV-2 were also positive, raising questions about a potential contribution. Against this, we found no metagenomic evidence for SARS-CoV-2 infection in liver or blood, while a larger UK series of 44 cases including the five transplants described here, found levels of SARS-CoV-2 infection and seropositivity to be to be 28% and 38% respectively^12^. The results of this case series are supported by preliminary data from a UKHSA study which has not shown differences in the prevalence of SARS-CoV-2 antibody between cases and age matched controls (UKHSA personal communication). In line with UK national recommendations at the time, none of the children had received a COVID vaccine.

While there is limited evidence for SARS-CoV-2 directly causing the hepatitis outbreak, the findings may also reflect the impact of the COVID-19 pandemic on child mixing and infection patterns. The contemporaneous development of unexplained hepatitis with an outbreak of HAdV-F41^2^and the finding of HAdV-F41 in many cases, suggests that the two are linked. Enteric adenovirus infection is most common in those aged under five^2^ and infection is influenced by mixing and hygiene^26^. Few cases of HAdV-F41 occurred between 2020 and 2022 and no major outbreaks were recorded ^2^. The current outbreak followed relaxation of restrictions due to the pandemic and represented one of many infections, including other enteric pathogens such as norovirus, that occurred in UK children following return to normal mixing^27^. Notwithstanding, the most consistent biomarker for the hepatitis of unknown origin is AAV2 which is present in 94% (16/17) of cases overall, and at high viral loads in 91% (10/11) of the cases with blood testing. In contrast, AAV2 was present less commonly and at much lower titres in controls selected on the basis of HAdV infection and/or hepatitis, specifically in 6% of 100 immunocompetent and 31% of 32 immunosuppressed controls. The one hepatitis case fulfilling the UKHSA definition, who was negative for AAV2, was also negative for HAdV by metagenomics and PCR (**Table 2**). With little information about this case, it is plausible that the absence of both HAdV and AAV2 could reflect loss of virus between onset of hepatitis and testing. Alternatively, there remains the possibility that it was misclassified. AAV2 is known to be spread with respiratory adenoviruses but has not been detected by us in over 30 SARS-CoV-2 positive nasopharyngeal aspirates (data not shown). Under normal circumstances, AAV-2 antibodies levels are high at birth, subsequently declining to reach their lowest point at 7-11 months, thereafter increasing through childhood and adolescence^27^. With loss of child mixing during the COVID pandemic, reduced spread of common respiratory and enteric viral infections and no evidence of transmission of AAV2 infection in association with SARS-CoV-2, it is likely that immunity to both HAdV-F41 and AAV2 declined sharply in the age group affected by this unexplained hepatitis outbreak. Although not previously described, we found AAV2 to be present in stool from both HAdV-F41 positive cases and controls. Taken together, it is likely that many children in the age cohort at peak risk of the unexplained hepatitis lacked immunity to both HAdV-F41 and AAV2 and may have experienced primary infection with both simultaneously.

In the absence of laboratory evidence for viral proteins or virion particles in liver biopsies, it is hard to conclude that cases of unexplained hepatitis were caused by direct lytic infection involving either or both viruses. HLA typing of 20 Scottish cases and 64 controls identified an association of the Class II HLA DRB1*0401 allele with cases ^28^, with lower odds ratios found for association with the DQA1*03:03 and DQB1*03:01 alleles. Analysis of the five transplanted cases in our series, found at least one of these alleles to be present in each of the five cases, with HLA DRB1*0401 being present in 4/5 (**Supplementary Table 8**). The proteomic data from two explanted livers also showed high levels of HLA Class II proteins. Together with the lack of evidence for lytic viral infection, these data provide further support for the possibility that the unexplained hepatitis is immune-mediated, occurring in children who are genetically predisposed albeit potentially triggered by infection with HAdV-F41 and/or AAV2.

There are a number of limitations to our study. First, our numbers are small and the finding is of an association of AAV2 presence and high viral levels with the unexplained paediatric hepatitis cases. While there is strong circumstantial evidence, we cannot establish causality with the data we have. AAV2 could be a bystander or a marker of co-infection with HAdV-F41s or even HHV-6B. Against this explanation is the association of high levels of AAV2 with cases of unexplained hepatitis and the much less frequent and statistically lower levels of AAV2 in all controls including those with hepatitis of known aetiology, even when infected with HAdV or even HHV-6 helper viruses. The relevance of this finding together with the potential contribution of AAV2 to this outbreak of unexplained hepatitis need further testing in properly controlled studies. Second, due to the low viral titres of adenovirus in whole blood and other specimens from the cases, only one full HAdV genome could be sequenced from the stool of a case. While this cannot completely exclude the emergence of a new HAdV-F41 as the cause of this outbreak, the clustering of this sequence with other UK HAdV-F41 sequences and the partial whole blood AdF41 sequences from cases, reduce the likelihood. Third, our data alone are not sufficient to rule out an effect of SARS-CoV-2 Omicron, the appearance of which preceded the outbreak of unexplained hepatitis and this will need to wait for results from large case-controlled studies such as the one underway led by UKHSA. Fourth, while we postulate that changes in the epidemiology of infections in children during the pandemic may have altered patterns of AAV2 and HAdV infection and disease, further data, with wider testing of other cohorts and seroepidemiological studies of AAV2 in cases and controls are needed to support this.

In summary, we find through metagenomic discovery and other methods that high levels of Adeno-associated virus 2 is a biomarker for cases of unexplained hepatitis occurring in children <5 years of age in the UK since January 2022 and this may even extend to cases of HAdV hepatitis occurring in immunocompromised children. While HAdV-F41 infection is also found, we hypothesise that it is a necessary but not sufficient prerequisite for development of the unexplained hepatitis. HAdV-F41 infections were widespread including in many children without hepatitis. We find little evidence to support direct lytic viral infection in explanted livers from cases by either HAdV-F41 or AAV2, with no viral proteins detected by either IHC or proteomic analysis. Instead, we find evidence for high expression of Class II HLA alleles in transplanted cases that have been postulated, in parallel studies to confer increased genetic risk for the unexplained hepatitis in these children. We hypothesise that, as a consequence of the disruption in childhood mixing patterns, resulting from the pandemic restrictions, young children may not have previously been exposed to HAdV-F41 or AAV2. The recent HAdV-F41 outbreaks that followed lifting of restrictions may also have resulted in AAV2 infection, with either or both viruses triggering an immune-mediated hepatitis in genetically susceptible children. Further studies are now needed to investigate this hypothesis. The putative association of AAV2 with liver disease in children, revealed by the use of powerful new metagenomic tools, also requires investigation in other cases of unexplained hepatitis.

## METHODS

### Ethics

Metagenomic analysis and adenovirus sequencing were carried out by the routine diagnostic service at Great Ormond Street Hospital. Additional PCRs, Immunohistochemistry and proteomics on samples received for metagenomics are part of the Great Ormond Street Hospital (GOSH) protocol for confirmation of new and unexpected pathogens. The use for research of anonymised laboratory request data, diagnostic results and residual material from any specimen received in the GOSH diagnostic laboratory, including all cases received from Birmingham’s Children Hospital UKHSA, Public Health Wales, Public health Scotland as well as non-case samples from UKHSA, Public Health Scotland and Great Ormond Street Hospital research was approved by UCL Partners Pathogen Biobank under ethical approval granted by the NRES Committee London-Fulham (REC reference: 17/LO/1530). Children undergoing liver transplant were consented for additional research under the International Severe Acute Respiratory and Emerging Infections Con Ethics sortium (ISARIC) WHO Clinical Characterisation Protocol UK (CCP-UK) [ISRCTN 66726260] (RQ3001-0591, RQ301-0594, RQ301-0596, RQ301-0597, RQ301-0598). Ethical approval for the ISARIC CCP-UK study was given by the South Central–Oxford C Research Ethics Committee in England (13/SC/0149), the Scotland A Research Ethics Committee (20/SS/0028), and the WHO Ethics Review Committee (RPC571 and RPC572).

The United Kingdom Health Security Agency (UKHSA) has legal permission, provided by Regulation 3 of The Health Service (Control of Patient Information) Regulations 2002, to process patient confidential information for national surveillance of communicable diseases and as such, individual patient consent is not required.

Control patients for DIAMONDS/PERFORM were recruited according to the approved enrolment procedures of each study, and with the informed consent of parents or guardians: DIAMONDS (London – Dulwich Research Ethics Committee: 20/HRA/1714); PERFORM (London – Central Research Ethics Committee: 16/LO/1684).

The sample IDs use for the cases are anonymised IDs that cannot reveal the identity of the study subjects and not known to anyone outside the research group, such as the patients or the hospital staff.

### Samples

Initial diagnostic testing by metagenomics and PCR was performed at Great Ormond Street Hospital Microbiology and Virology clinical laboratories. Further whole genome sequencing and characterization was performed at UCL.

### Case samples from Birmingham Children’s Hospital, UKHSA, Public Health Wales and Public Health Scotland / Edinburgh Royal Infirmary

We received explanted liver tissue from five biopsy sites from five cases, five whole blood 500ul from four cases and serum plasma from one case (**Table 1, Figure 1B**). These were used in metagenomics testing (**Table 2A**), followed by HAdV, HHV-6 and AAV2 testing by PCR and, depending on CT value, whole genome sequencing (**Table 2B, Supplementary Table 3-5**). We subsequently received 25 additional specimens from UKHSA, Public Health Wales and Public Health Scotland / Edinburgh Royal Infirmary, including 16 additional blood samples, 4 respiratory specimens and 5 stool samples, for HAdV WGS and depending on residual material for AAV2 PCR testing followed by sequencing (**Table 1, Table 2, Figure 1B, Supplementary Table 3-5**).

### Control Samples from GOSH for AAV2 and HHV-6 PCR

Blood samples from 32 patients not linked to the non-A-E hepatitis outbreak were tested by real-time PCR targeting AAV2, as detailed in Table 3. These were purified DNA from residual diagnostic specimens received in the GOSH Microbiology and Virology laboratory in the previous year. All residual specimens were stored at -80 °C prior to testing and pseudo-anonymised at the point of processing and analysis.

Twenty-six controls were patients with ALT/AST >500 and Adenovirus, HHV-6, CMV or EBV viraemia. Viraemia was initially detected using targeted real-time PCR during routine diagnostic testing with UKAS-accredited lab-developed assays that conform to ISO:15189 standards.

Five controls were patients with SARS-CoV2-positive nose and throat swabs, for whom residual blood samples were available. SARS-CoV2 PCR was not performed on the blood samples. One additional blood sample was from a patient with MIS-C syndrome.

In addition to the blood samples, four residual liver biopsies from four control patients referred for investigation of infection were tested by AAV2 and HHV-6 PCR. The liver biopsies were submitted to the GOSH microbiology laboratory for routine diagnosis by bacterial broad-range 16S rRNA gene PCR or metagenomics testing in 2021 and 2022. 3/4 of the control patients were known to have elevated liver enzymes.

### Control samples from DIAMONDS and PERFORM

PERFORM (Personalised Risk assessment in Febrile illness to Optimise Real-life Management across the European Union) recruited children from 10 EU countries (2016-2020. PERFORM was funded by the European Union’s Horizon 2020 program under GA No 668303.

DIAMONDS (Diagnosis and Management of Febrile Illness using RNA Personalised Molecular Signature Diagnosis) is funded by the European Union Horizon 2020 program grant number 848196. Recruitment commenced in 2020, and is ongoing. Both studies recruited children presenting with suspected infection or inflammation, and assigned them to diagnostic groups according to a standardised algorithm.

### Control samples from UKHSA

We received a blood sample from one patient with raised liver enzymes and HAdV infection. We also received one control stool sample from Public Health Scotland/Edinburgh Royal Infirmary and 22 control stool samples for sequencing.

### Metagenomic sequencing

#### Nucleic acid purification for metagenomics

Frozen liver biopsies were infused overnight at -20°C with RNAlater-ICE. Up to 20 mg biopsy was lysed with 1.4mm ceramic, 0.1mm silica and 4mm glass beads, prior to DNA and RNA purification using the Qiagen AllPrep DNA/RNA Mini kit as per manufacturers’ instructions, with a 30 µl elution volume for RNA and 50 µl for DNA.

Up to 400 µl whole blood was lysed with 0.5mm and 0.1 mm glass beads prior to DNA and RNA purification on a Qiagen EZ1 instrument with an EZ1 virus mini kit as per manufacturer’s instructions, with a 60 µl elution volume.

For quality assurance, every batch of samples was accompanied by a control sample containing feline calicivirus RNA and cowpox DNA which was processed alongside clinical specimens, from nucleic acid purification through to sequencing. All specimens and controls were spiked with MS2 phage RNA internal control prior to nucleic acid purification.

#### Metagenomics library preparation and sequencing

RNA from whole blood samples with RNA yield >2.5 ng/µl and from biopsies underwent ribosomal RNA depletion and library prep with KAPA RNA HyperPrep kit with RiboErase, according to manufacturer’s instructions. RNA from whole blood with RNA yield <2.5 ng/µl did not undergo rRNA depletion prior to library prep.

DNA from whole blood samples with DNA yield >1 ng/µl and from biopsies underwent depletion of CpG-methylated DNA using the NEBNext® Microbiome DNA Enrichment Kit, followed by library preparation with NEBNext Ultra II FS DNA Library Prep Kit for Illumina, according to manufacturer’s instructions. DNA from whole blood with DNA yield <1 ng/µl did not undergo depletion of CpG-methylated DNA prior to library prep.

Sequencing was performed with a NextSeq High output 150 cycle kit with a maximum of 12 libraries pooled per run, including controls.

#### Metagenomics data analysis

##### Pre-processing pipeline

An initial quality control step was performed by trimming adapters and low-quality ends from the reads (TrimGalore - https://www.bioinformatics.babraham.ac.uk/projects/trim_galore/, version 0.3.7). Human sequences were then removed using the human reference GRCH38 p.9 (Bowtie2, version 2.4.1 ^29^) followed by removal of low quality and low complexity sequences (PrinSeq, version 0.20.3^30^). An additional step of human seq removal followed (megaBLAST, version 2.9.0). For RNA-seq, ribosomal RNA sequences were also removed using a similar 2 step-approach (Bowtie2 and megaBLAST). Finally, nucleotide similarity and protein similarity searches were performed (megaBLAST and DIAMOND^31^ (version 0.9.30) respectively) against custom reference databases that consisted of nucleotide and protein sequences of the RefSeq collections (downloaded March 2020) for viruses, bacteria, fungi, parasites and human.

##### Taxonomic classification

DNA and RNA sequence data was analysed with metaMix^32^ (version 0.4) nucleotide and protein analysis pipelines.

metaMix resolves metagenomics mixtures using Bayesian mixture models and parallel MCMC search of the potential species space to infer the most likely species profile.

metaMix considers all reads simultaneously to infer relative abundances and probabilistically assign the reads to the species most likely to be present. It uses an ‘unknown’ category to capture the fact that some reads cannot be assigned to any species. The resulting metagenomic profile includes posterior probabilities of species presence as well as Bayes factor for presence versus absence of specific species. There are two modes, metaMix-protein, which is optimal for RNA virus detection and metaMix-nucl, which is best for speciation of DNA microbes. Both modes were used for RNA-seq while metaMix-nucl for DNA-seq.

For sequence results to be valid, MS2 phage RNA had to be detected in every sample and feline calicivirus RNA and cowpox DNA, with no additional unexpected organisms, detected in the controls.

##### Confirmatory mapping of AAV2

The RNA-Seq reads were mapped to the AAV2 reference genome (NCBI reference sequence NC_001401) using Bowtie2, with the –very-sensitive option. Samtools^33^ version 1.9) and Picard (version 2.26.9, http://broadinstitute.github.io/picard/) were used to sort, deduplicate and index the alignments, and to create a depth file, which was plotted using a custom script in R.

##### *de novo* assembly of unclassified reads

We performed a *de novo* assembly step with metaSPADES^34^(v3.15.5), using all the reads with no matches to the nucleotide database we used for our similarity search. A search using megaBLAST with the standard nucleotide collection was carried out on all resulting contigs over 1000bp in length.

### Proteomics

Patients’ tissues were homogenized in lysis buffer, 100 mM Tris (pH 8.5), 5% Sodium dodecyl sulfate, 5 mM tris(2-carboxyethyl)phosphine, 20 mM chloroacetamide then heated at 95 degrees for 10 minutes and sonicated in ultrasonic bath for other 10. The lysed proteins were quantified with NanoDrop 2000 (Thermo Fisher Scientific). 100 µg were precipitated with Methanol/Chloroform protocol and then protein pellets were reconstituted in 100 mM tris (pH 8.5) and 4% sodium deoxycholate (SDC). The proteins were subjected to proteolysis with 1:50 trypsin overnight at 37°C with constant shaking. Digestion was stopped by adding 1% trifluoroacetic acid to a final concentration of 0.5%. Precipitated SDC was removed by centrifugation at 10,000g for 5 min, and the supernatant containing digested peptides was desalted on an SOLAµ HRP (Thermo Fisher Scientific). Peptides were dried and dissolved in 2% formic acid before liquid chromatography–tandem mass spectrometry (MS/MS) analysis. A total of 2000 ng of the mixture of tryptic peptides was analysed using an Ultimate3000 high-performance liquid chromatography system coupled online to an Eclipse mass spectrometer (Thermo Fisher Scientific). Buffer A consisted of water acidified with 0.1% formic acid, while buffer B was 80% acetonitrile and 20% water with 0.1% formic acid. The peptides were first trapped for 1 min at 30 μl/min with 100% buffer A on a trap (0.3 mm by 5 mm with PepMap C18, 5 μm, 100 Å; Thermo Fisher Scientific); after trapping, the peptides were separated by a 50-cm analytical column (Acclaim PepMap, 3 μm; Thermo Fisher Scientific). The gradient was 9 to 35% B in 103 min at 300 nl/min. Buffer B was then raised to 55% in 2 min and increased to 99% for the cleaning step. Peptides were ionized using a spray voltage of 2.1 kV and a capillary heated at 280°C. The mass spectrometer was set to acquire full-scan MS spectra (350 to 1400 mass/charge ratio) for a maximum injection time set to Auto at a mass resolution of 120,000 and an automated gain control (AGC) target value of 100%. For a second the most intense precursor ions were selected for MS/MS. HCD fragmentation was performed in the HCD cell, with the readout in the Orbitrap mass analyser at a resolution of 15,000 (isolation window of 3 Th) and an AGC target value of 200% with a maximum injection time set to Auto and a normalized collision energy of 30%. All raw files were analysed by MaxQuant v2.1 software using the integrated Andromeda search engine and searched against the Human UniProt Reference Proteome (February release with 79,057 protein sequences) together with UniProt reported AAVs proteins and specific fasta created using EMBOSS Sixpack translating patient’s virus genome. MaxQuant^35^ was used with the standard parameters with only the addition of deamidation (N) as variable modification. Data analysis was then carried out with Perseus v2.05: Proteins reported in the file “proteinGroups.txt” were filtered for reverse and potential contaminants. Figures were created using Origin pro version 2022b.

### PCR

Real-time PCR targeting a 62 nt region of the AAV2 inverted terminal repeat (ITR) sequence was performed using primers and probes previously described ^36^. This assay is predicted to amplify AAV2 and AAV6. The Qiagen QuantiNova probe PCR kit (PERFORM and DIAMONDS controls) or Qiagen Quantifast probe PCR kit (all other samples) were used. Each 25 µl reaction consisted of 0.1 µM forward primer, 0.34 µM reverse primer, 0.1 µM probe with 5 µl template DNA.

Real-time PCR targeting a 74 bp region of the HHV-6 DNA polymerase gene was performed using primers and probes previously described ^37^ multiplexed with an internal positive control targeting mouse (*mus*) DNA spiked into each sample during DNA purification, as previously described^38^. Briefly, each 25 µl reaction consisted of 0.5 µM each HHV-6 primer, 0.3 µM HHV-6 probe, 0.12 µM each *mus* primer, 0.08 µM *mus* probe and 12.5 µl Qiagen Quantifast Fast mastermix with 10 µl template DNA.

Real-time PCR targeting a 132 bp region of the Adenovirus hexon gene was performed using primers and probes previously described ^39^ multiplexed with an internal positive control targeting mouse (*mus*) DNA spiked into each sample during DNA purification, as previously described (Tann *et al*. 2014). Briefly, each 25 µl reaction consisted of 0.6 µM each HHV-6 primer, 0.4 µM HHV-6 probe, 0.12 µM each *mus* primer, 0.08 µM *mus* probe and 12.5 µl Qiagen Quantifast Fast mastermix with 10 µl template DNA.

PCR cycling for all targets, apart from the controls from the PERFORM and DIAMONDS studies, was performed on an ABI 7500 Fast thermocycler and consisted of 95 °C for 5 minutes followed by 45 cycles of 95 °C for 30 seconds and 60 °C for 30 seconds. For the PERFORM and DIAMONDS controls, PCR was performed on a StepOnePlus™ Real-Time PCR System and consisted of 95 °C for 2 minutes followed by 45 cycles of 95 °C for 5 seconds and 60 °C for 10 seconds. Each PCR run included a no template control and a DNA positive control for each target.

### Immunohistochemistry (IHC)

All IHC was done on Formalin Fixed Paraffin Embedded tissue cut at 3mm thickness.

### Adenovirus

Adenovirus immunohistochemistry was carried out using the Ventana Benchmark ULTRA, Optiview Detection Kit, PIER with Protease 1 for 4min, Ab incubation 32min (Adenovirus clone 2/6 & 20/11, Roche, 760-4870, pre-diluted). The positive control was a known Adenovirus positive gastrointestinal surgical case.

### AAV2

#### Preparation of AAV2 positive controls

The plasmid used for transfection was pAAV2/2 (addgene, Plasmid #104963, https://www.addgene.org/104963/) which expresses the Rep/Cap genes of AAV2. This was delivered by tail-vein hydrodynamic injection ^40^into albino C57Bl/6 mice (5 microgrammes in 2 mls PBS). Negative ontrols received PBS alone. At 48 hours, mice were terminally exsanguinated and perfused by PBS. Livers were collected into 10% Neutral Buffered Formalin (CellPath UK). This was performed under Home Office License PAD4E6357.

##### AAV2 immunohistochemistry was carried out with three commercial kits

- Leica Bond-III, Bond Polymer Refine Detection Kit with DAB Enhancer, HIER with Bond Epitope Retrieval Solution 1 (citrate based pH 6) for 30min, Ab incubation 30min (Anti-AAV VP1/VP2/VP3 clone B1, PROGEN, 690058S, 1:100).
- Leica Bond-III, Bond Polymer Refine Detection Kit with DAB Enhancer, HIER with Bond Epitope Retrieval Solution 1 (citrate based pH 6) for 40min, Ab incubation 30min (Anti-AAV VP1/VP2/VP3 rabbit polyclonal, OriGene, BP5024, 1:100)

##### HHV-6 immunohistochemistry straining was carried out with

Leica Bond-III, Bond Polymer Refine Detection Kit with DAB Enhancer, PIER with Bond Enzyme 1 Kit 10min, Ab incubation 30min (Mouse monoclonal [C3108-103] to HHV-6, ABCAM, ab128404, 1:100).

### Electron Microscopy

Samples of liver were fixed in 2.5% glutaraldehyde in 0.1M cacodylate buffer followed by secondary fixation in 1.0% osmium tetroxide. Tissues were dehydrated in graded ethanol, transferred to an intermediate reagent, propylene oxide and then infiltrated and embedded in Agar 100 epoxy resin. Polymerisation was undertaken at 60 °C for 48 hours. 90nm ultrathin sections were cut using a Diatome diamond knife on a Leica UC7 ultramicrotome. Sections were transferred to copper grids and stained with alcoholic urynal acetate and Reynold’s lead citrate. The samples were examined using a JEOL 1400 transmission electron microscope. Images were captured on an AMT XR80 digital camera.

### Whole genome sequencing

#### Bait Design

Tio produce the capture probes for hybridisation, biotinylated RNA oligonucleotides (baits) used in the SureSelectXT protocols for HAdV and HHV-6 WGS were designed in-house using Agilent community design baits with part numbers 5191-6711 and 5191-6713 respectively. For AAV2 we designed amplicons based on an alignment of 18 complete genomes available from GenBank along with 1 sequence assembled from metagenomic analysis of the samples. Primers were designed using PrimalScheme v 1.3.2 with an average amplicon size of 400. A total of 15 primer pairs were designed as part of the scheme. They were synthesised by Agilent Technologies, Santa Clara, California (Agilent Technologies, 2021) (available through Agilent’s Community Designs programme: SSXT CD Pan Adenovirus and SSXT CD Pan HHV-6 and used previously ^41,42^).

#### Library prep and sequencing

For whole genome sequencing of HAdV and HHV-6, DNA (bulked with male human gDNA (Promega) if required) was sheared using a Covaris E220 focused ultra-sonication system (PIP 75, duty factor 10, cycles per burst 1000).End-repair, non-templated addition of 3′ poly A, adapter ligation, hybridisation, PCR (pre-capture cycles dependent on DNA input and post capture cycles dependent on viral load), and all post-reaction clean-up steps were performed according to either the SureSelectXT Low Input Target Enrichment for Illumina Paired-End Multiplexed Sequencing protocol (version A0), the SureSelectXT Target Enrichment for Illumina Paired-End Multiplexed Sequencing protocol (version C3)or SureSelectXTHS Target Enrichment using the Magnis NGS Prep System protocol (version A0) (Agilent Technologies). Quality control steps were performed on the 4200 TapeStation (Agilent Technologies). Samples were sequenced using the Illumina MiSeq platform. Base calling and sample demultiplexing were performed as standard for the MiSeq platform, generating paired FASTQ files for each sample. A negative control was included on each processing run.

For WGS of AAV-2: an AAV-2 primer scheme was designed using primalscheme^43^ with 17 AAV-2 sequences from NCBI and 1 AAV-2 sequence provided by GOSH from metagenomic sequencing of a liver biopsy DNA extract as the reference material. These primers amplify 15 overlapping 400 bp amplicons. Primers were supplied by Merck. Two multiplex PCR reactions were prepared using Q5® Hot Start High-Fidelity 2X Master Mix, with a 65°C, 3 min annealing/extension temperature. Pool 1 and 2 multiplex PCRs were run for 35 cycles.10uL of each PCR reaction were combined and 20uL nuclease-free water added. Libraries were prepared either manually or on the Agilent Bravo NGS workstation option B, following a reduced-scale version of the Illumina DNA protocol as used in the CoronaHiT protocol^44^. Equal volumes of the final libraries were pooled, bead purified and sequenced on the Illumina MiSeq. A negative control was included on each processing run.

All library preps and sequencing were performed by UCL Genomics.

### AAV2 Sequence Analysis

The raw fastq reads were adapted, trimmed and low-quality reads removed. The reads were mapped to NC_001401 reference sequence and then the amplicon primers regions were trimmed using the location provided in a bed file. Consensus sequences were then called at a minimum of 10X coverage. The entire processing of raw reads to consensus was carried out using nf-core/viralrecon pipeline (https://nf-co.re/viralrecon/2.4.1) (doi:https://doi.org/10.5281/zenodo.3901628). Basic quality metrics for the samples sequenced are in Supplementary Table 5. All samples that gave 10x genome coverage over 90% were then used for further phylogenetic analysis. Samples were aligned along with known reference strains from genbank using MAFFT (version v7.271) and the trees were built with IQ-TREE (multicore version 1.6.12) with 1000 rapid bootstraps and aLRT support. The samples were then labelled based on type and provider on the trees (Fig 3A).

For each AAV2 sample, we aligned the consensus nucleotide sequence to the AAV2 reference sequence. From these alignments, the exact coordinates of the sample capsid were determined. We then used the coordinates to extract the corresponding nucleotide sequence and translated it to find the amino acid sequence. We then compared each sample to the reference to identify amino acid changes. Amino acid sequences from AAV capsid sequences were retrieved from GenBank for AAV1 to AAV12. Amino acid sequences of capsid constructs designed to be more hepatotropic were retrieved from ^24,25^. These sequence sets were then aligned to the AAV2 reference sequence using MAFFT^45^. We then compared each construct to the AAV2 reference to identify amino acid changes present, while retaining the AAV2 coordinate set.

### HAdV and HHV-6 sequence analysis

Raw data quality control is performed using trim-galore (v.0.6.7) on the raw FASTQ files.

For HHV-6, short reads were mapped with BWA mem^46^ (0.7.17-r1188) using the RefSeq reference NC_000898.

For adenovirus, genotyping is performed using AYUKA (version 22-111, Guerra-Assuncao, manuscript in preparation). This novel tool is used to confidently assign one or more adenovirus genotypes to a sample of interest, assessing inter-genotype recombination if more than one genotype detected. The results from this screening step guide which downstream analyses are performed, and which reference genome(s) are used. If mixed infection is suspected, reads are separated using bbsplit (https://sourceforge.net/projects/bbmap/), and each genotype is analysed independently as normal. If recombination is suspected, a more detailed analysis is performed using RDP and the sample is excluded from phylogenetic analysis. After genotyping, the cleaned read data is mapped using BWA to the relevant reference sequence(s), single nucleotide polymorphisms and small insertions and deletions are called using bcftool (version1.15.1, https://github.com/samtools/bcftools) and a consensus sequence is generated also with bcftools, masking with Ns positions that do not have enough read support (15X by default). Consensus sequences generated with the pipeline are then concatenated to previously sequenced samples and a multiple sequence alignment is performed using the G-INS-I algorithm in the MAFFT software (MAFFT G-INS-I v7.481). The multiple sequence alignment is then used for phylogenetic analysis with IQ-TREE^47^ (IQ-TREE 2 2.2.0), using modelfinder and performing 1000 rapid bootstraps.

### Statistical Analysis

Fisher’s exact test and two-sided Wilcoxon (Mann-Whitney) non-parametric rank sum test were used for differences between case and control groups. Where multiple groups were compared, Kruskal-Wallis tests followed by Wilcoxon pairwise tests using a Benjamini-Hochberg correction were performed. All analysis were performed in R version 4.2.0.

## Supporting information

Consortia authors

## Data Availability

All data produced in the present study are available upon reasonable request to the authors

## Data and Code Availability

The consensus genomes from WGS data are currently being deposited in Genbank. Available upon request

## Acknowledgements

UKHSA for funding of the metagenomics and adenovirus sequencing.

All research at Great Ormond Street Hospital NHS Foundation Trust and UCL Great Ormond Street Institute of Child Health is made possible by the NIHR Great Ormond Street Hospital Biomedical Research Centre. The views expressed are those of the author(s) and not necessarily those of the NHS, the NIHR or the Department of Health.

## Supplementary Figures

**Supplementary Figure 1:**
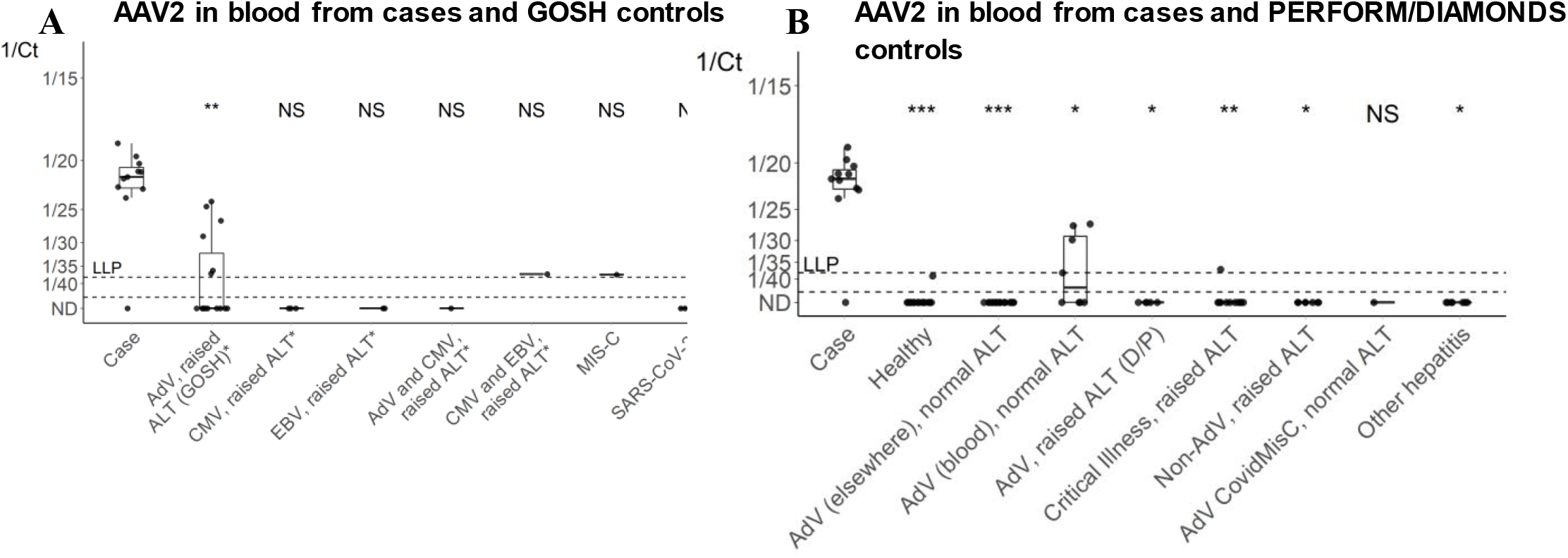
viral loads (CT values) for AAV2 controls under 10 years old. Case control comparison with control patients under 10 years of age only. See Figure 2 legend for more details.

**Supplementary Figure 2:**
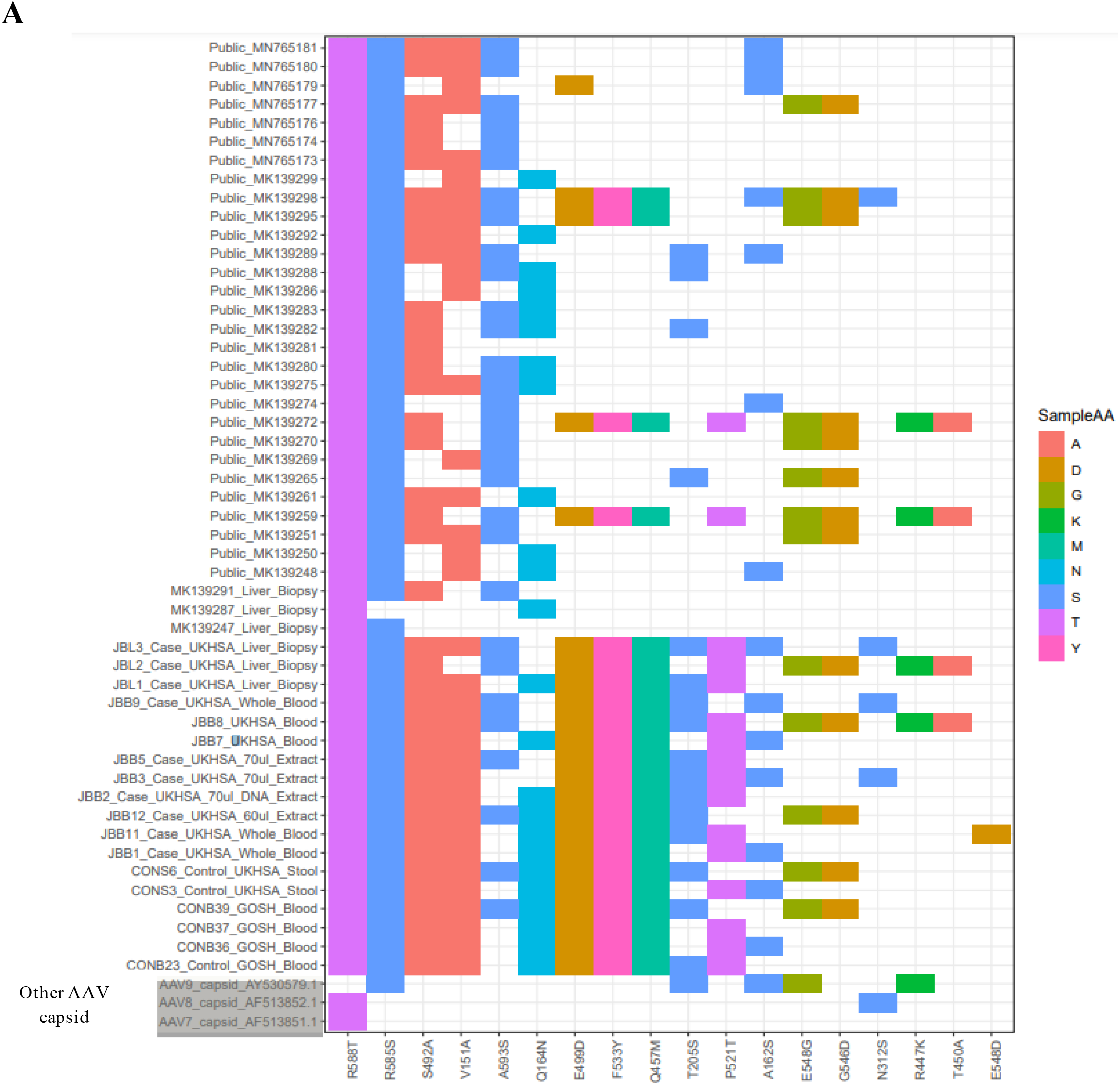
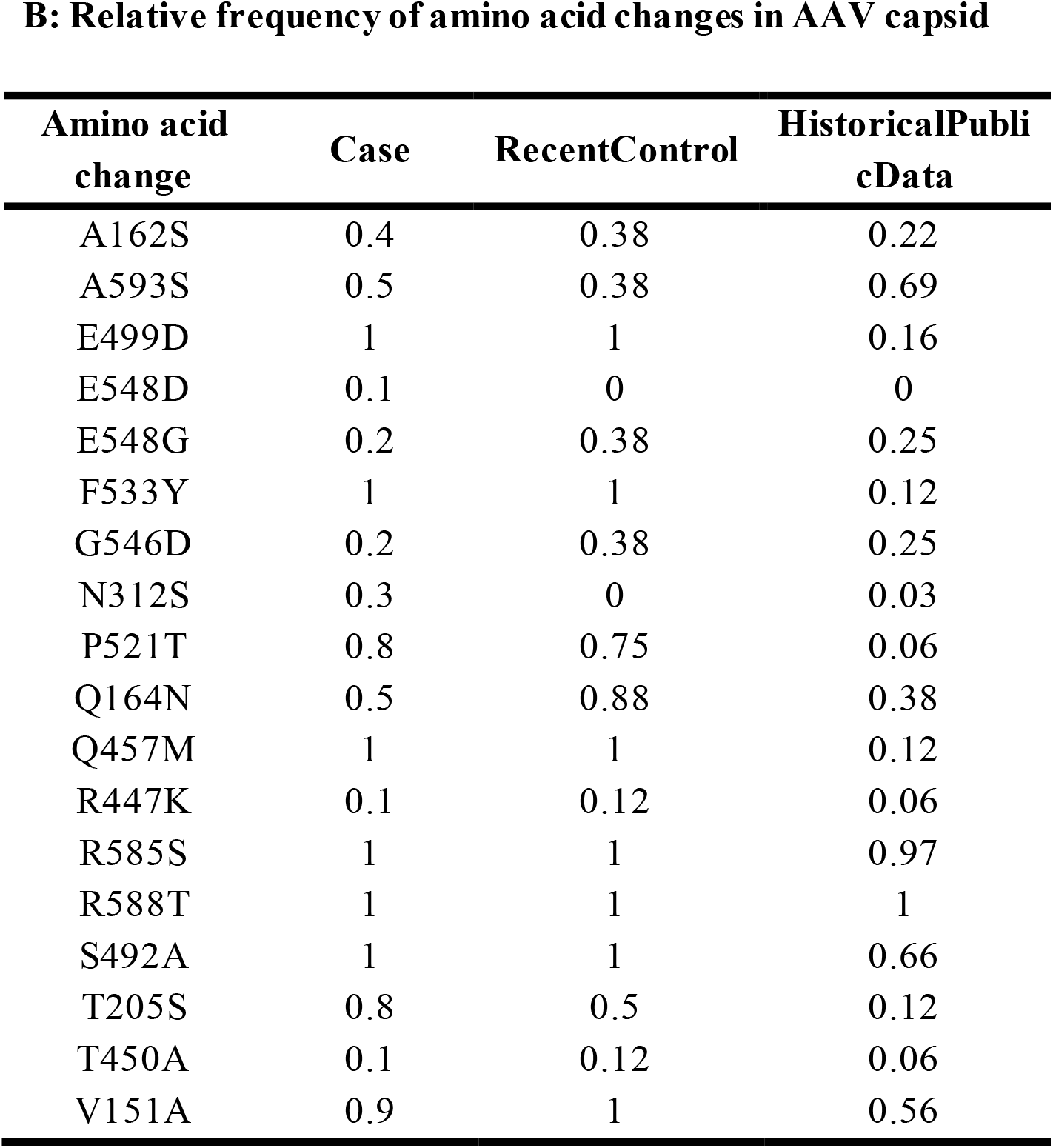
AAV2 and HHV-6 sequence analysis. A) Amino acid differences between AAV2 capsid sequences from cases, contemporaneously circulating controls and historical publicly available sequences compared with the and the AVV2 reference sequence NC_001401.2. Also shown are the capsid sequences from known AAV7,8 and 9 hepatotropic capsids compared to the reference sequence NC_001401.2. B) Frequency table of capsid residues in cases and historical controls. There is no difference between the capsid sequences of cases and contemporaneously circulating controls. However, there are changes compared with historical controls in all contemporary sequences. None of the recently acquired capsid changes are shared with known hepatotrophic strains in AAV7, 8 and 9.

**Supplementary Figure 3:**
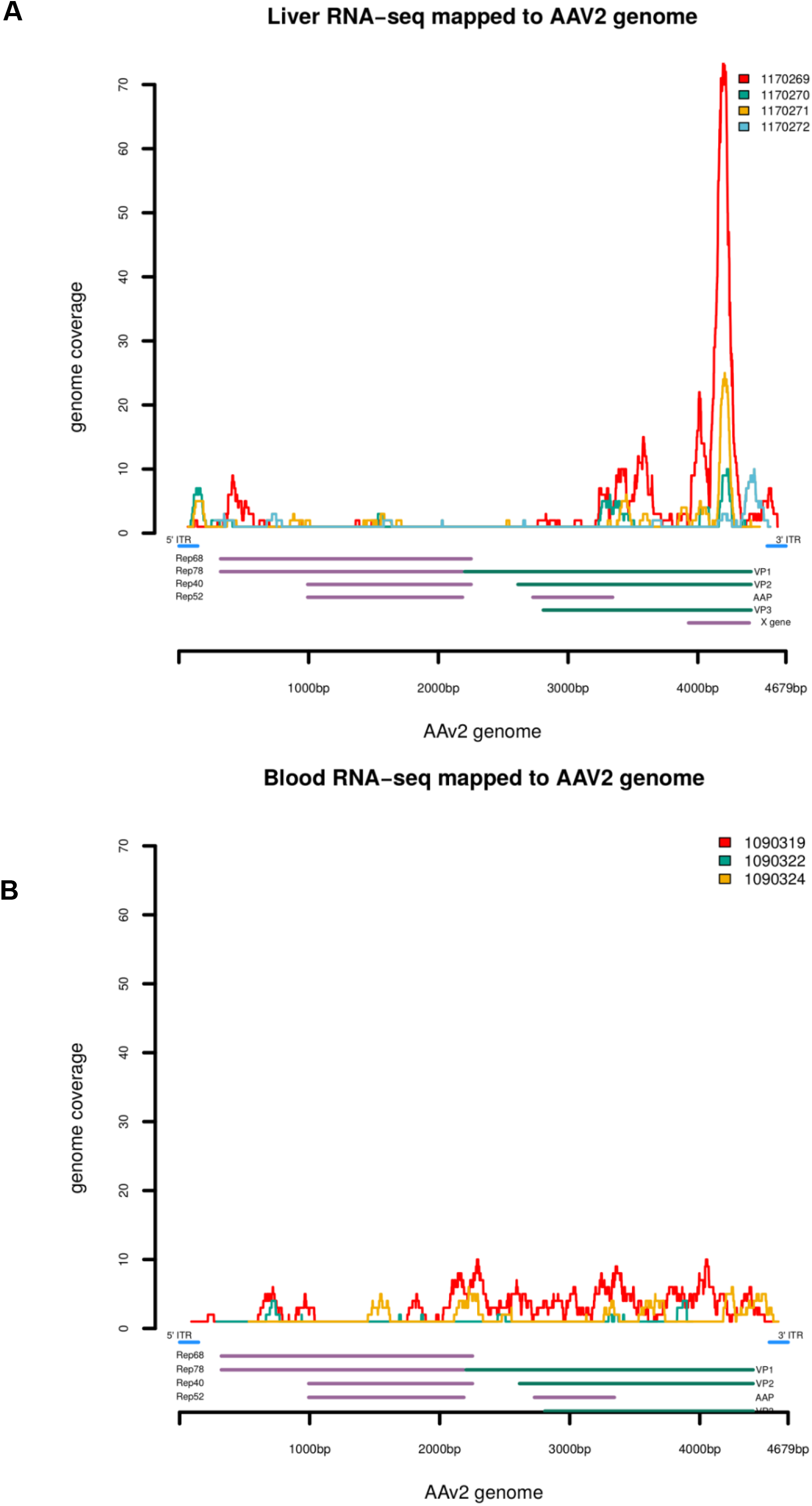
Evidence of AAV2 replication from meta-transcriptomics. Mapping of AAV2 reads to the reference genome for A) Liver RNA-Seq from 4 cases, B) blood RNA-Seq from 3 cases

**Supplementary Figure 4:**
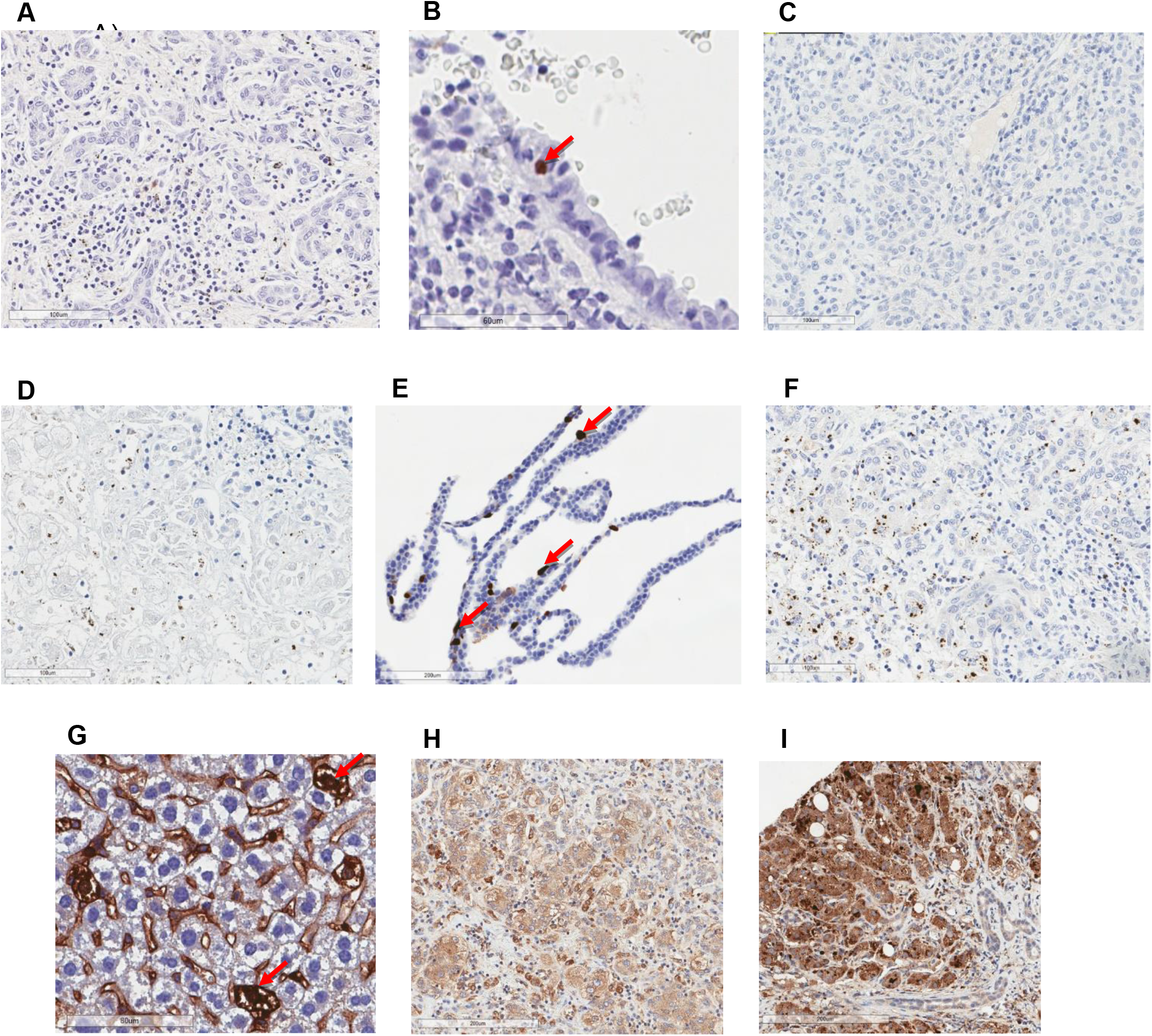
Histopathology. Immunohistochemical (IHC) staining for HAdV, AAV2 and HHV-6 on cases and controls. Amouse monoclonal antibody as used for A) Adenovirus IHC on a case, B) Adenovirus IHC on a control, C) AAV2 IHC on case 3a, D) AAV2 IHC on case 3b, E) AAV2 IHC on a control.A rabbit polyclonal antibody was used for F) AAV2 IHC on case 2, G) positive control, H) HHV-6 IHC on case 4, I) HHV-6 on a control. Examples of positively stained control cells are indicated by arrows. Clumps of cytoplasmic positivity in occasional residual hepatocytes, are non-specific and interpreted as likely to be phagocytosed debris.

**Supplementary Figure 5:**
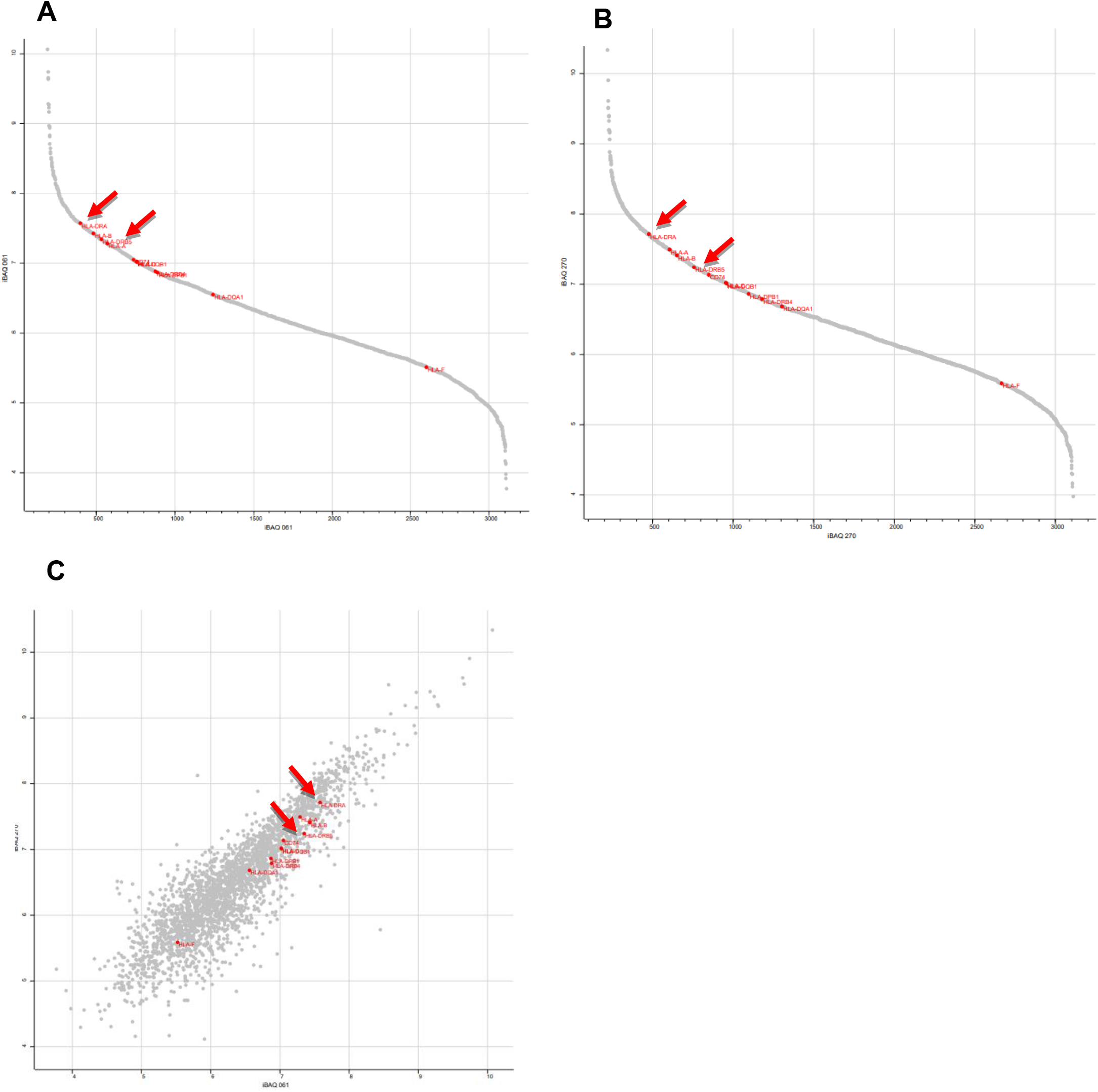
Proteomics. A-B) Ranking of the quantified proteins using the log10 of iBAQ values for A) JBL4 and B) JBL5. C) Scatter plot of quantified proteins in sample JBL4 versus JBL5. HLA proteins are highlighted in red. Red arrows denote HLA-DRA and HLA-DRB.

## Supplementary Tables

**Supplementary Table 1:** Clinical details for transplanted patients. Median age 3y (range:1y-4y).

| Patient | Sex | Ethnicity | Symptoms | presenting WBC | AST | ALT PEAK | GGT | Albumin |
| --- | --- | --- | --- | --- | --- | --- | --- | --- |
| **JBL4** | M | caucasian | J,V,D,PALE | 10 |  | 3564 | 126 | 36 |
| **JBL2** | M | caucasian | J,P,V,D,PALE | nk | 7510 | 2927 | 59 | 31 |
| **JBL1** | M | caucasian | J, V, D, F | 11 | 4750 | 2819 | 165 | 34 |
| **JBL3** | F | caucasian | J, V | 12 | 4511 | 4742 | 119 | 36 |
| **JBL5** | F | caucasian | V,C,D,J | 6.8 | 3637 | 3576 | nk | 30 |

| Patient | Presenting PT | PT PEA K | Hep A/B/C/E | EB V | CMV | Adeno throat | Adeno stool | Adeno 1 |
| --- | --- | --- | --- | --- | --- | --- | --- | --- |
| **JBL4** | 16 | 36 | neg | neg | neg | nt | neg | 7908 |
| **JBL2** | 15 | 45 | neg | neg | neg | pos | nt | 29594 |
| **JBL1** | 17 | 55 | neg | neg | neg | pos | pos | 11951 |
| **JBL3** | 15 | 37 | neg | neg | neg | nt | neg | 29741 |
| **JBL5** | 20 | 47 | neg | neg | neg | nt | nt | 25419 |

| Patient | Covid Ab | Covid PCR | IgG | Ferritin | Other viruses | Symptom to TX in days |
| --- | --- | --- | --- | --- | --- | --- |
| **JBL4** | neg | pos | 11.6 | 594 | HHV-6, Entero, Lepto, HSV,VZV, neg | 12 |
| **JBL2** | nt | pos | nt | nt | HHV-6 pos | 8 |
| **JBL1** | nt | pos | nt | 4605 | HHV-6 pos, HHV-7 neg | 12 |
| **JBL3** | nt | neg | 10.8 | 233 | HHV-6 pos, HHV-7 neg | 14 |
| **JBL5** | pos | neg | nt | nt | FluA/B, RSV neg, parvovirus neg | 18 |

PT: prothrombin peak, AST: aspartate aminotransferase, ALT: alanine aminotransferase, GGT: gamm glutamyl transferase
J: jaundice, V: vomiting, D: diarrhoea, PALE: pale stool, F: fever, C: coryza, P: abdominal pain
Pos: positive, neg: negative, nt: not tested, nk: not known

**Supplementary Table 2:** Metagenomics summary statistics: raw read counts, human filtered and other findings.

| Specim | RNA |  |  | DNA |  |  |
| --- | --- | --- | --- | --- | --- | --- |
|  | Raw reads | Human filtered | Other mNGS results | Raw reads | Human filtered | Other mNGS results |
| **Liver** |  |  |  |  |  |  |
| JBL1 | 57400446 | 895604 | - | 32273777 | 1250914 | Astrovirus VA3, TTV |
| JBL4 | 63430844 | 1028780 | - | 49625805 | 2584180 | - |
| JBL3 | 63430844 | 1093392 | - | 44881648 | 1542178 | TTV |
| JBL2 | 66916975 | 879266 | - | 49660375 | 1611202 | - |
| JBL5 | F | F | - | 86643490 | TBC | - |
| **Blood** |  |  |  |  |  |  |
| JBPL1 | 21053703 | 117728 | TTV | 21131740 | 293772 | TTV |
| JBB14 | 13615796 | 89174 | - | 24110121 | 318254 | TTV |
| JBB16 | 11394711 | 67128 | - | 56728541 | 794514 | TTV |
| JBB10 | F | F | F | 38172955 | 357120 | EBV |
| JBB1 | 50388499 | 357810 | - | 46101012 | 482796 | TTV |
| JBB15 | F | F | F | 44186067 | 459964 | - |

**Supplementary Table 3:**
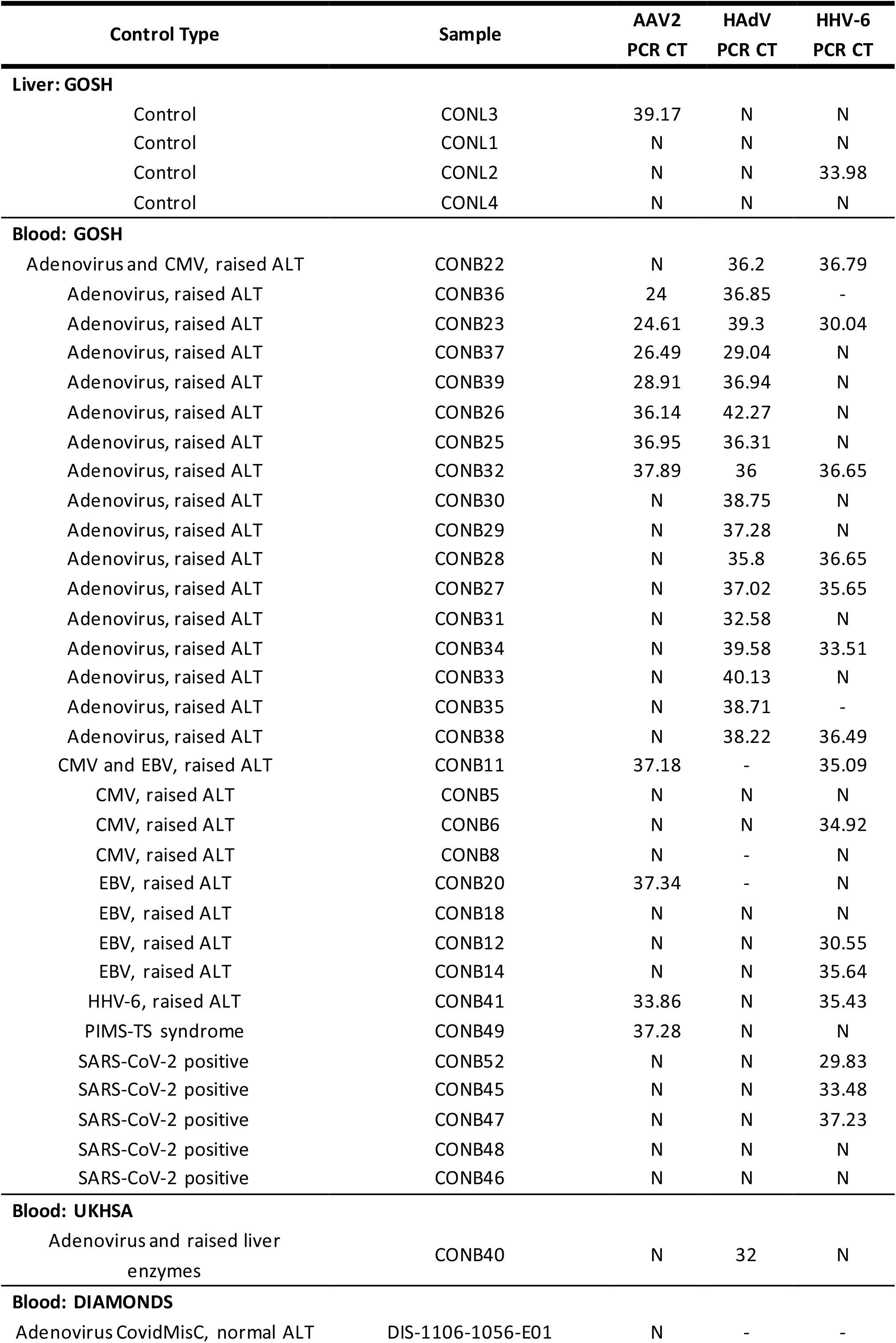
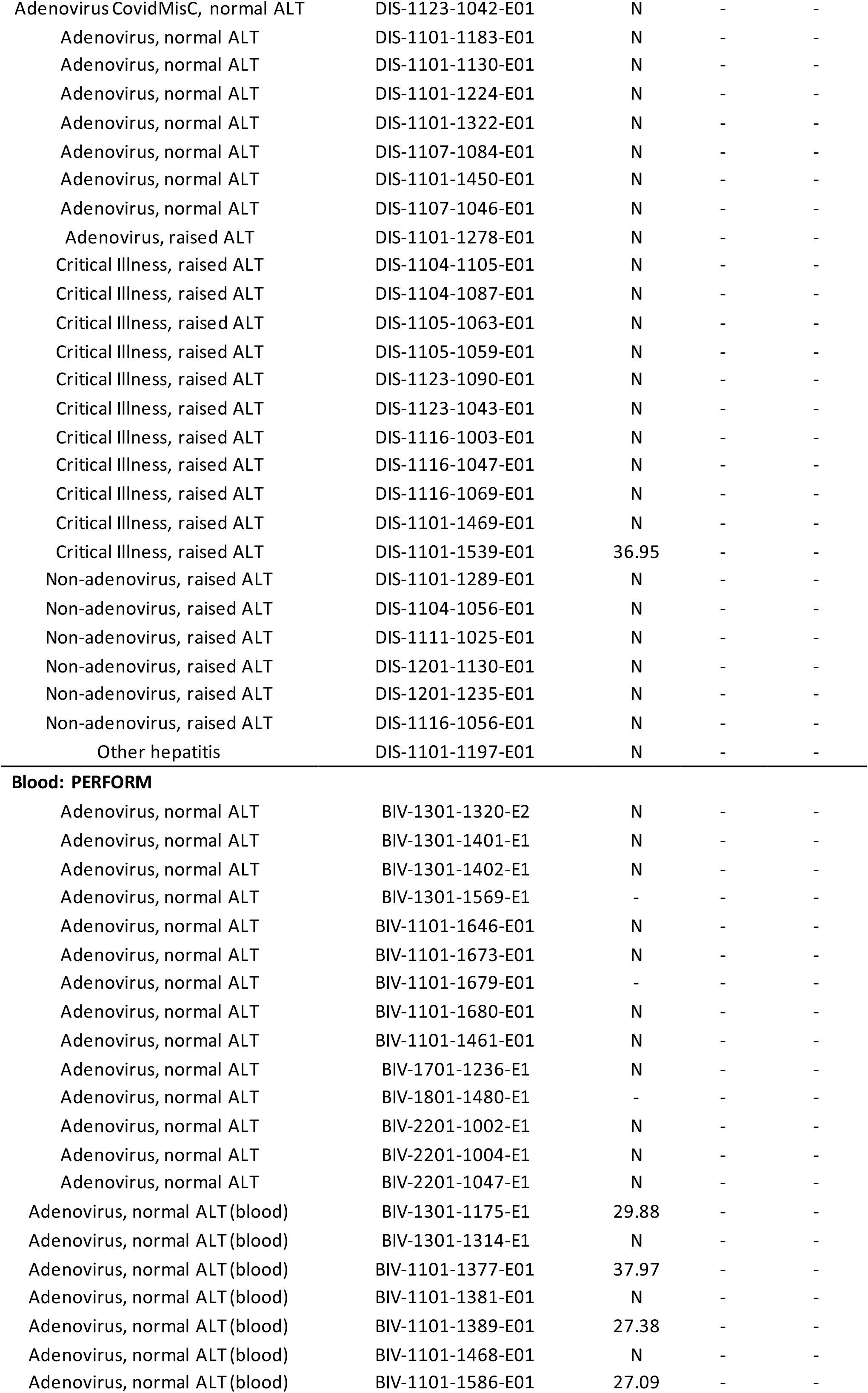
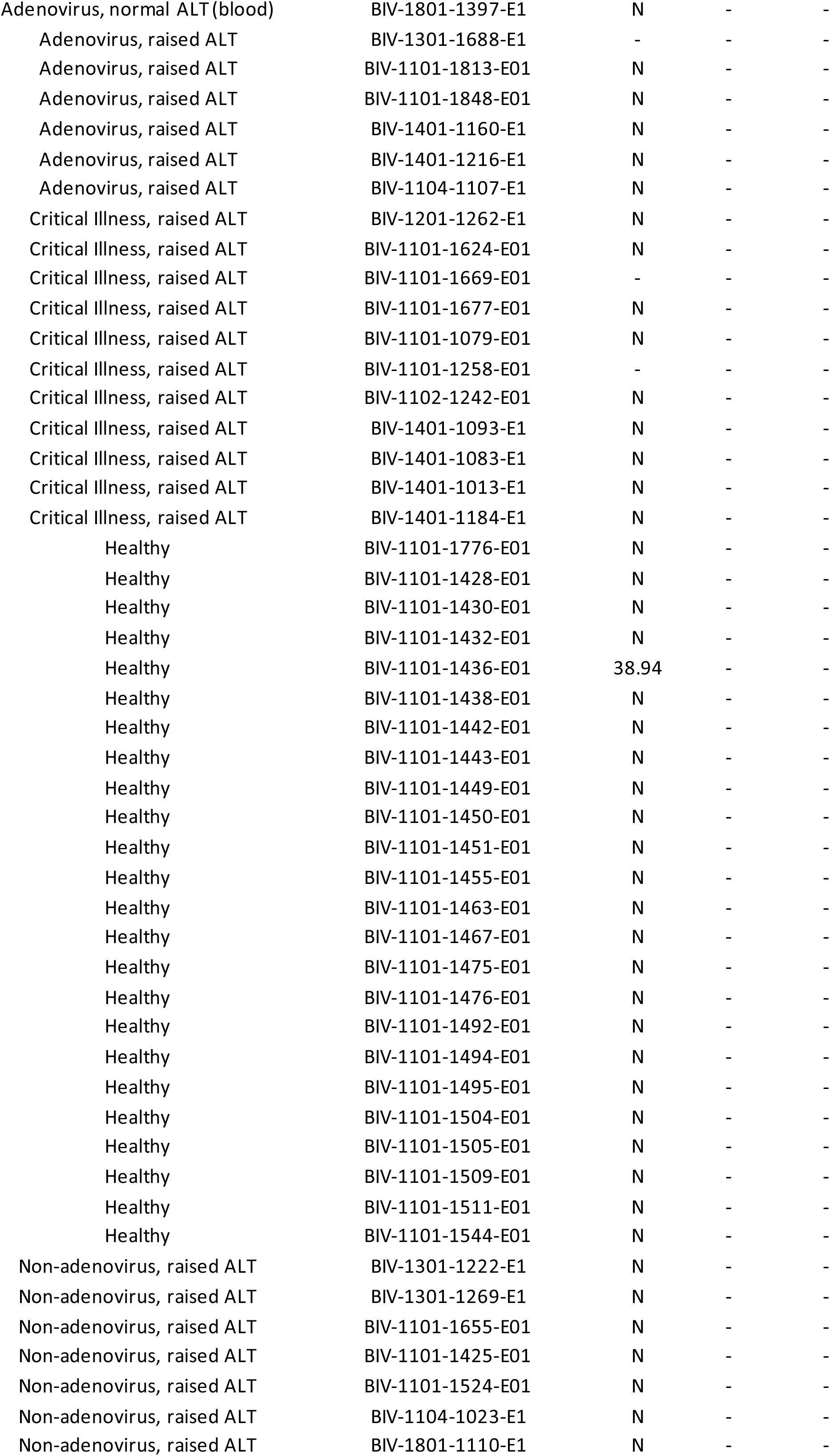
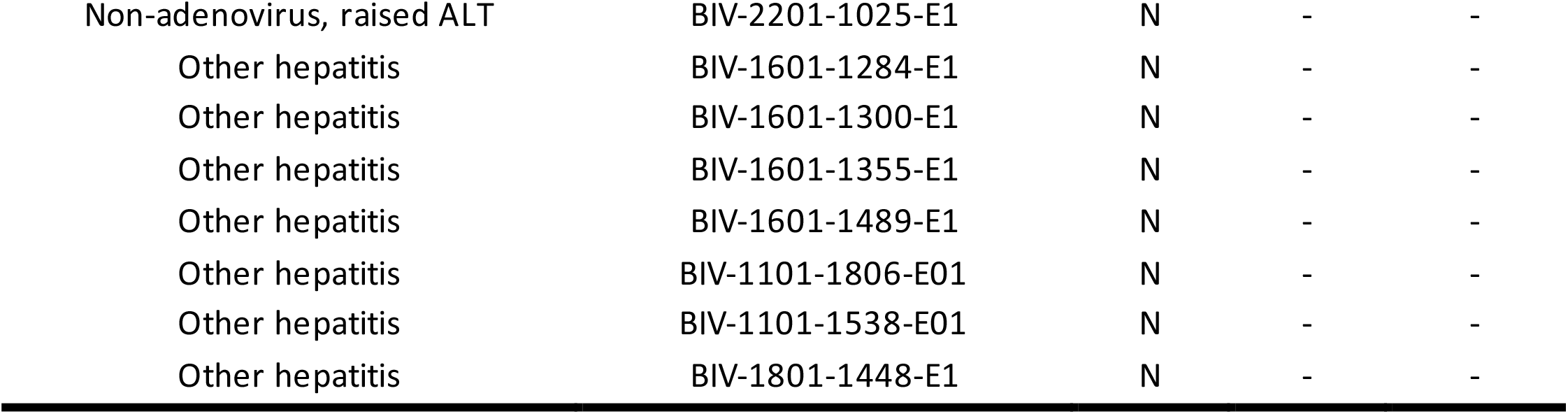
Control PCR Results.

| Control Type | Sample | AAV2<br>PCR CT | HAdV<br>PCR CT | HHV-6<br>PCR CT |
| --- | --- | --- | --- | --- |
| **Liver: GOSH** |  |  |  |  |
| Control | CONL3 | 39.17 | N | N |
| Control | CONL1 | N | N | N |
| Control | CONL2 | N | N | 33.98 |
| Control | CONL4 | N | N | N |
| **Blood: GOSH** |  |  |  |  |
| Adenovirus and CMV, raised ALT | CONB22 | N | 36.2 | 36.79 |
| Adenovirus, raised ALT | CONB36 | 24 | 36.85 | - |
| Adenovirus, raised ALT | CONB23 | 24.61 | 39.3 | 30.04 |
| Adenovirus, raised ALT | CONB37 | 26.49 | 29.04 | N |
| Adenovirus, raised ALT | CONB39 | 28.91 | 36.94 | N |
| Adenovirus, raised ALT | CONB26 | 36.14 | 42.27 | N |
| Adenovirus, raised ALT | CONB25 | 36.95 | 36.31 | N |
| Adenovirus, raised ALT | CONB32 | 37.89 | 36 | 36.65 |
| Adenovirus, raised ALT | CONB30 | N | 38.75 | N |
| Adenovirus, raised ALT | CONB29 | N | 37.28 | N |
| Adenovirus, raised ALT | CONB28 | N | 35.8 | 36.65 |
| Adenovirus, raised ALT | CONB27 | N | 37.02 | 35.65 |
| Adenovirus, raised ALT | CONB31 | N | 32.58 | N |
| Adenovirus, raised ALT | CONB34 | N | 39.58 | 33.51 |
| Adenovirus, raised ALT | CONB33 | N | 40.13 | N |
| Adenovirus, raised ALT | CONB35 | N | 38.71 | - |
| Adenovirus, raised ALT | CONB38 | N | 38.22 | 36.49 |
| CMV and EBV, raised ALT | CONB11 | 37.18 | - | 35.09 |
| CMV, raised ALT | CONB5 | N | N | N |
| CMV, raised ALT | CONB6 | N | N | 34.92 |
| CMV, raised ALT | CONB8 | N | - | N |
| EBV, raised ALT | CONB20 | 37.34 | - | N |
| EBV, raised ALT | CONB18 | N | N | N |
| EBV, raised ALT | CONB12 | N | N | 30.55 |
| EBV, raised ALT | CONB14 | N | N | 35.64 |
| HHV-6, raised ALT | CONB41 | 33.86 | N | 35.43 |
| PIMS-TS syndrome | CONB49 | 37.28 | N | N |
| SARS-CoV-2 positive | CONB52 | N | N | 29.83 |
| SARS-CoV-2 positive | CONB45 | N | N | 33.48 |
| SARS-CoV-2 positive | CONB47 | N | N | 37.23 |
| SARS-CoV-2 positive | CONB48 | N | N | N |
| SARS-CoV-2 positive | CONB46 | N | N | N |
| **Blood: UKHSA** |  |  |  |  |
| Adenovirus and raised liver enzymes | CONB40 | N | 32 | N |
| **Blood: DIAMONDS** |  |  |  |  |
| Adenovirus CovidMisC, normal ALT | DIS-1106-1056-E01 | N | - | - |
| Adenovirus CovidMisC, normal ALT | DIS-1123-1042-E01 | N | - | - |
| Adenovirus, normal ALT | DIS-1101-1183-E01 | N | - | - |
| Adenovirus, normal ALT | DIS-1101-1130-E01 | N | - | - |
| Adenovirus, normal ALT | DIS-1101-1224-E01 | N | - | - |
| Adenovirus, normal ALT | DIS-1101-1322-E01 | N | - | - |
| Adenovirus, normal ALT | DIS-1107-1084-E01 | N | - | - |
| Adenovirus, normal ALT | DIS-1101-1450-E01 | N | - | - |
| Adenovirus, normal ALT | DIS-1107-1046-E01 | N | - | - |
| Adenovirus, raised ALT | DIS-1101-1278-E01 | N | - | - |
| Critical Illness, raised ALT | DIS-1104-1105-E01 | N | - | - |
| Critical Illness, raised ALT | DIS-1104-1087-E01 | N | - | - |
| Critical Illness, raised ALT | DIS-1105-1063-E01 | N | - | - |
| Critical Illness, raised ALT | DIS-1105-1059-E01 | N | - | - |
| Critical Illness, raised ALT | DIS-1123-1090-E01 | N | - | - |
| Critical Illness, raised ALT | DIS-1123-1043-E01 | N | - | - |
| Critical Illness, raised ALT | DIS-1116-1003-E01 | N | - | - |
| Critical Illness, raised ALT | DIS-1116-1047-E01 | N | - | - |
| Critical Illness, raised ALT | DIS-1116-1069-E01 | N | - | - |
| Critical Illness, raised ALT | DIS-1101-1469-E01 | N | - | - |
| Critical Illness, raised ALT | DIS-1101-1539-E01 | 36.95 | - | - |
| Non-adenovirus, raised ALT | DIS-1101-1289-E01 | N | - | - |
| Non-adenovirus, raised ALT | DIS-1104-1056-E01 | N | - | - |
| Non-adenovirus, raised ALT | DIS-1111-1025-E01 | N | - | - |
| Non-adenovirus, raised ALT | DIS-1201-1130-E01 | N | - | - |
| Non-adenovirus, raised ALT | DIS-1201-1235-E01 | N | - | - |
| Non-adenovirus, raised ALT | DIS-1116-1056-E01 | N | - | - |
| Other hepatitis | DIS-1101-1197-E01 | N | - | - |
| **Blood: PERFORM** |  |  |  |  |
| Adenovirus, normal ALT | BIV-1301-1320-E2 | N | - | - |
| Adenovirus, normal ALT | BIV-1301-1401-E1 | N | - | - |
| Adenovirus, normal ALT | BIV-1301-1402-E1 | N | - | - |
| Adenovirus, normal ALT | BIV-1301-1569-E1 | - | - | - |
| Adenovirus, normal ALT | BIV-1101-1646-E01 | N | - | - |
| Adenovirus, normal ALT | BIV-1101-1673-E01 | N | - | - |
| Adenovirus, normal ALT | BIV-1101-1679-E01 | - | - | - |
| Adenovirus, normal ALT | BIV-1101-1680-E01 | N | - | - |
| Adenovirus, normal ALT | BIV-1101-1461-E01 | N | - | - |
| Adenovirus, normal ALT | BIV-1701-1236-E1 | N | - | - |
| Adenovirus, normal ALT | BIV-1801-1480-E1 | - | - | - |
| Adenovirus, normal ALT | BIV-2201-1002-E1 | N | - | - |
| Adenovirus, normal ALT | BIV-2201-1004-E1 | N | - | - |
| Adenovirus, normal ALT | BIV-2201-1047-E1 | N | - | - |
| Adenovirus, normal ALT (blood) | BIV-1301-1175-E1 | 29.88 | - | - |
| Adenovirus, normal ALT (blood) | BIV-1301-1314-E1 | N | - | - |
| Adenovirus, normal ALT (blood) | BIV-1101-1377-E01 | 37.97 | - | - |
| Adenovirus, normal ALT (blood) | BIV-1101-1381-E01 | N | - | - |
| Adenovirus, normal ALT (blood) | BIV-1101-1389-E01 | 27.38 | - | - |
| Adenovirus, normal ALT (blood) | BIV-1101-1468-E01 | N | - | - |
| Adenovirus, normal ALT (blood) | BIV-1101-1586-E01 | 27.09 | - | - |
| Adenovirus, normal ALT (blood) | BIV-1801-1397-E1 | N | - | - |
| Adenovirus, raised ALT | BIV-1301-1688-E1 | - | - | - |
| Adenovirus, raised ALT | BIV-1101-1813-E01 | N | - | - |
| Adenovirus, raised ALT | BIV-1101-1848-E01 | N | - | - |
| Adenovirus, raised ALT | BIV-1401-1160-E1 | N | - | - |
| Adenovirus, raised ALT | BIV-1401-1216-E1 | N | - | - |
| Adenovirus, raised ALT | BIV-1104-1107-E1 | N | - | - |
| Critical Illness, raised ALT | BIV-1201-1262-E1 | N | - | - |
| Critical Illness, raised ALT | BIV-1101-1624-E01 | N | - | - |
| Critical Illness, raised ALT | BIV-1101-1669-E01 | - | - | - |
| Critical Illness, raised ALT | BIV-1101-1677-E01 | N | - | - |
| Critical Illness, raised ALT | BIV-1101-1079-E01 | N | - | - |
| Critical Illness, raised ALT | BIV-1101-1258-E01 | - | - | - |
| Critical Illness, raised ALT | BIV-1102-1242-E01 | N | - | - |
| Critical Illness, raised ALT | BIV-1401-1093-E1 | N | - | - |
| Critical Illness, raised ALT | BIV-1401-1083-E1 | N | - | - |
| Critical Illness, raised ALT | BIV-1401-1013-E1 | N | - | - |
| Critical Illness, raised ALT | BIV-1401-1184-E1 | N | - | - |
| Healthy | BIV-1101-1776-E01 | N | - | - |
| Healthy | BIV-1101-1428-E01 | N | - | - |
| Healthy | BIV-1101-1430-E01 | N | - | - |
| Healthy | BIV-1101-1432-E01 | N | - | - |
| Healthy | BIV-1101-1436-E01 | 38.94 | - | - |
| Healthy | BIV-1101-1438-E01 | N | - | - |
| Healthy | BIV-1101-1442-E01 | N | - | - |
| Healthy | BIV-1101-1443-E01 | N | - | - |
| Healthy | BIV-1101-1449-E01 | N | - | - |
| Healthy | BIV-1101-1450-E01 | N | - | - |
| Healthy | BIV-1101-1451-E01 | N | - | - |
| Healthy | BIV-1101-1455-E01 | N | - | - |
| Healthy | BIV-1101-1463-E01 | N | - | - |
| Healthy | BIV-1101-1467-E01 | N | - | - |
| Healthy | BIV-1101-1475-E01 | N | - | - |
| Healthy | BIV-1101-1476-E01 | N | - | - |
| Healthy | BIV-1101-1492-E01 | N | - | - |
| Healthy | BIV-1101-1494-E01 | N | - | - |
| Healthy | BIV-1101-1495-E01 | N | - | - |
| Healthy | BIV-1101-1504-E01 | N | - | - |
| Healthy | BIV-1101-1505-E01 | N | - | - |
| Healthy | BIV-1101-1509-E01 | N | - | - |
| Healthy | BIV-1101-1511-E01 | N | - | - |
| Healthy | BIV-1101-1544-E01 | N | - | - |
| Non-adenovirus, raised ALT | BIV-1301-1222-E1 | N | - | - |
| Non-adenovirus, raised ALT | BIV-1301-1269-E1 | N | - | - |
| Non-adenovirus, raised ALT | BIV-1101-1655-E01 | N | - | - |
| Non-adenovirus, raised ALT | BIV-1101-1425-E01 | N | - | - |
| Non-adenovirus, raised ALT | BIV-1101-1524-E01 | N | - | - |
| Non-adenovirus, raised ALT | BIV-1104-1023-E1 | N | - | - |
| Non-adenovirus, raised ALT | BIV-1801-1110-E1 | N | - | - |
| Non-adenovirus, raised ALT | BIV-2201-1025-E1 | N | - | - |
| Other hepatitis | BIV-1601-1284-E1 | N | - | - |
| Other hepatitis | BIV-1601-1300-E1 | N | - | - |
| Other hepatitis | BIV-1601-1355-E1 | N | - | - |
| Other hepatitis | BIV-1601-1489-E1 | N | - | - |
| Other hepatitis | BIV-1101-1806-E01 | N | - | - |
| Other hepatitis | BIV-1101-1538-E01 | N | - | - |
| Other hepatitis | BIV-1801-1448-E1 | N | - | - |

**Supplementary Table 4:**
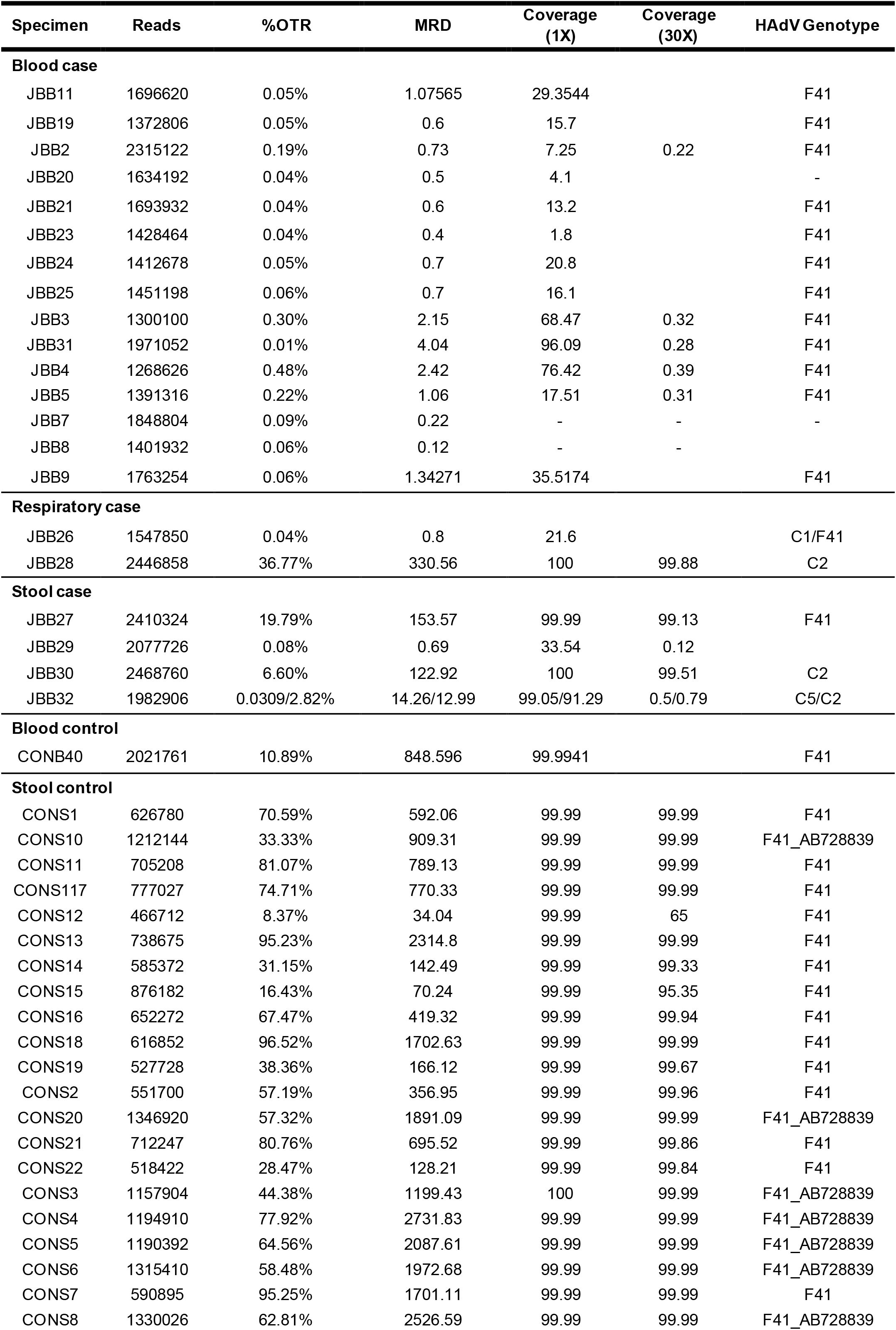
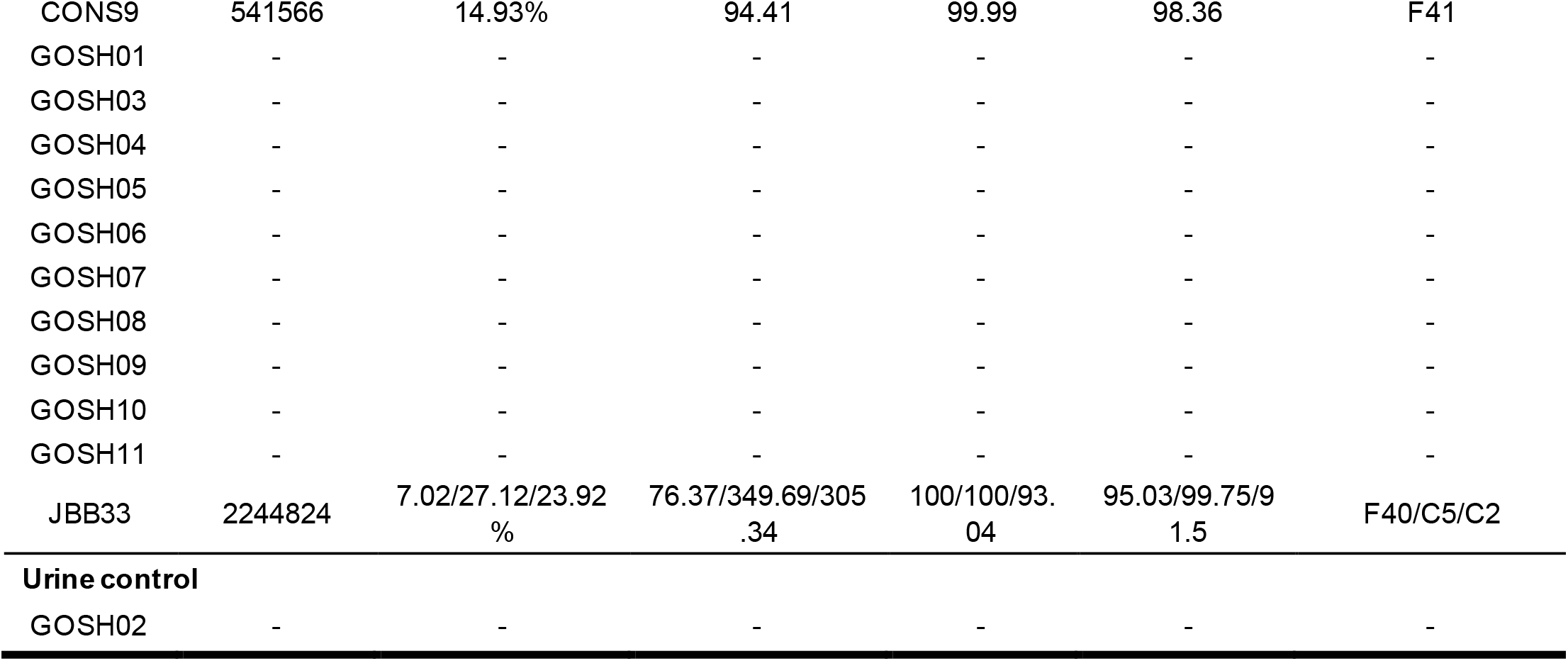
HAdV Whole Genome Sequencing.

**Supplementary Table 5:**
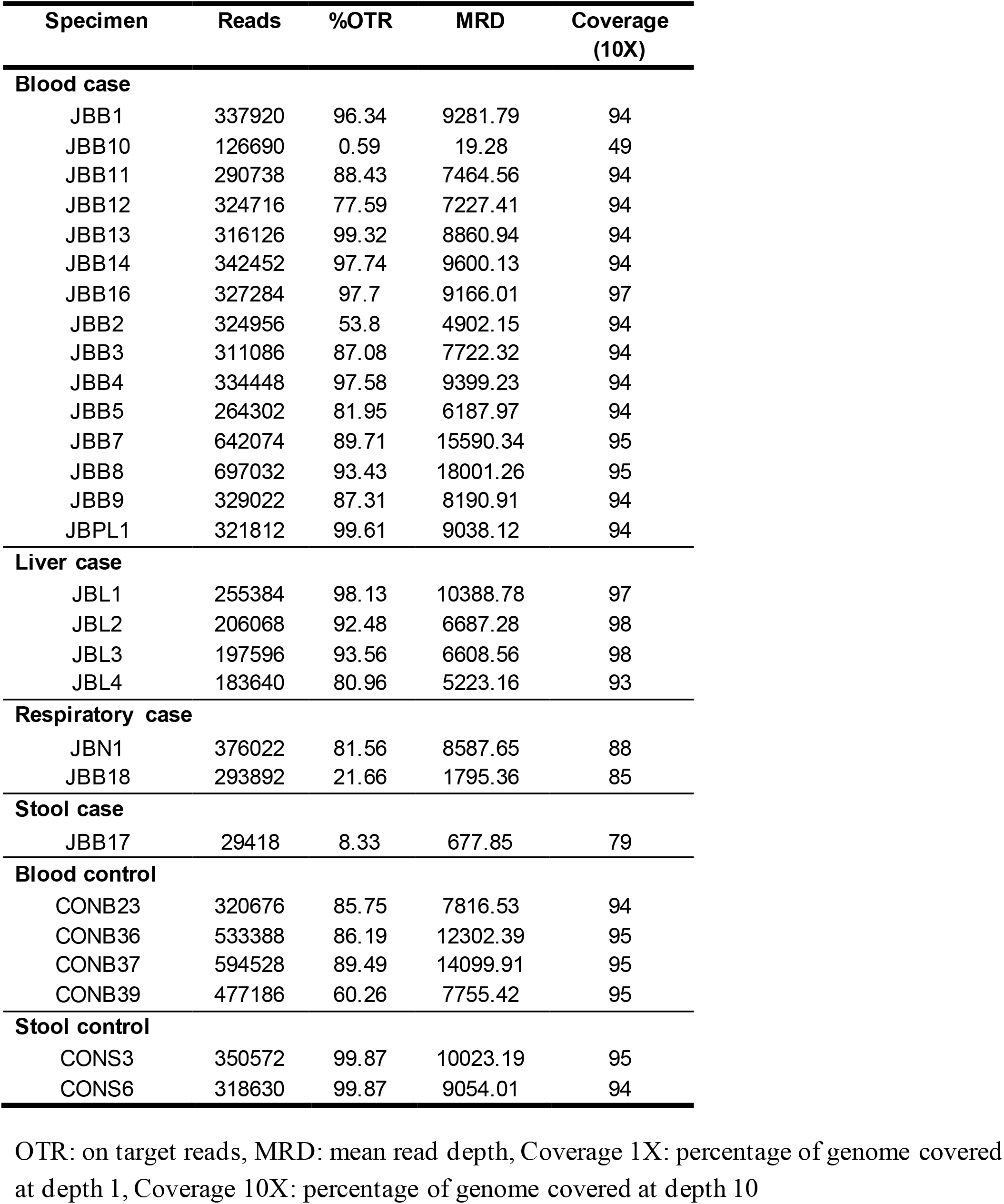
AAV2 Sequencing.

**Supplementary Table 6:** HHV-6 Sequencing of liver case samples.

| **Sample** | **Total Reads** | **OTR** | **MRD** | **Coverage 1X** | **Coverage 10X** |
| --- | --- | --- | --- | --- | --- |
| JBL1 | 1309913 | 9.82% | 76.43 | 93.89 | 2.67 |
| JBL4 | 1409364 | 10.25% | 91.45 | 91.7 | 2.06 |
| JBL3 | 1469731 | 10.46% | 98.06 | 87.5 | 1.9 |
| JBL2 | 1274320 | 16.73% | 193.84 | 96.58 | 93.58 |

OTR: on target reads, MRD: mean read depth, Coverage 1X: percentage of genome covered at depth 1, Coverage 10X: percentage of genome covered at depth 10

**Supplementary Table 7:**
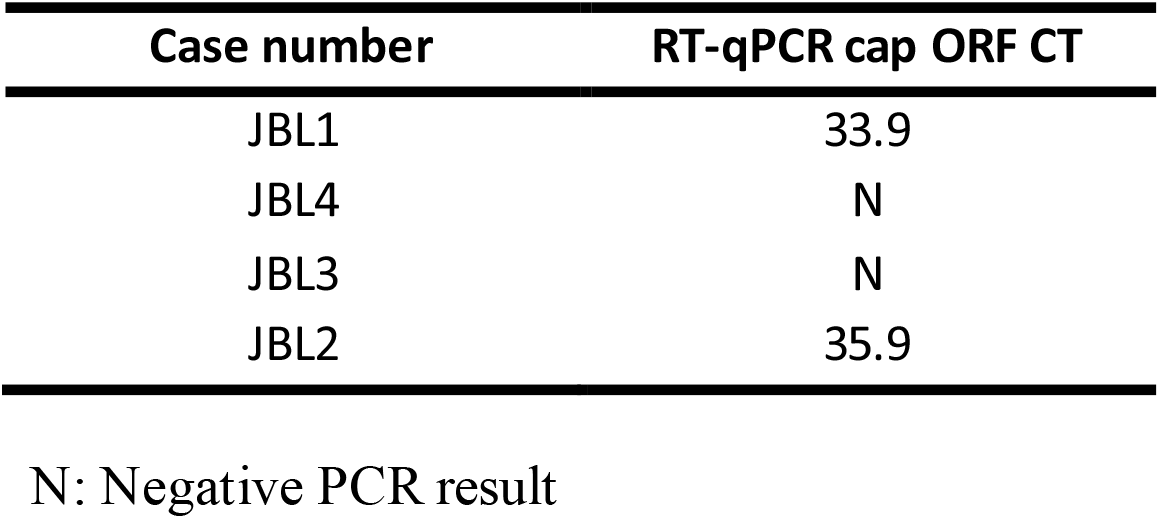
RT-PCR results for liver cases.

| **Case number** | **RT-qPCR cap ORF CT** |
| --- | --- |
| JBL1 | 33.9 |
| JBL4 | N |
| JBL3 | N |
| JBL2 | 35.9 |

N: Negative PCR result

**Supplementary Table 8:** HLA alleles identified as over-represented in Scottish hepatitis cases.

| **Patient** | **Number of HLA Alleles** |  |  |
| --- | --- | --- | --- |
|  | **DRB1\*04:01** | **DQA1\*03:03** | **DQB1\*03:01** |
| JBL1 | 1 | 2 | 2 |
| JBL2 | 0 | 1 | 1 |
| JBL3 | 2 | 1 | 1 |
| JBL4 | 1 | 0 | 0 |
| JBL5 | 2 | 2 | 1 |

